# Spatial mapping of Ethiopian cutaneous leishmaniasis lesions reveals distinct tissue level immune programs

**DOI:** 10.64898/2026.02.04.26345554

**Authors:** Nidhi S. Dey, Thao-Thy Pham, Shoumit Dey, Mekibib Kassa, Tigist Mekonnen, Helina Fikre, Pieter Monsieurs, SPATIAL CL CONSORTIUM, Myrthe Pareyn, Johan van Griensven, Malgorzata Domagalska, Jean-Claude Dujardin, Mezgebu Silamsaw Asres, Mikias Woldetensay, Paul M. Kaye, Wim Adriaensen

## Abstract

Ethiopian cutaneous leishmaniasis (CL) shows remarkably heterogeneous clinical presentations, but the underlying immunopathological mechanisms driving this heterogeneity in disease presentation remains poorly understood. To characterise the local immune response in Ethiopian CL, we performed spatial transcriptomics on paired lesional and non-lesional skin punch biopsies from five Ethiopian CL patients. We used reference-free deconvolution, morphology-guided regional analyses, and immunohistochemistry to identify five spatially distinct immunopathological responses that could co-exist within individual lesions: i) epithelial hyperplasia with interferon-stimulated keratinocytes, ii) cytotoxicity with tertiary lymphoid structures, iii) granulomatous inflammation with proinflammatory response iv) granulomatous inflammation with M2-polarised myeloid cell responses, and v) fibrotic remodelling with active collagen synthesis. Future longitudinal studies are needed to distinguish whether these patterns represent discrete pathotypes or stages of disease progression and to enable immune-based patient stratification and precision therapeutic interventions.

## Introduction

The cutaneous leishmaniases (CL), caused by infection with several different species of *Leishmania*, are some of the most important neglected tropical diseases affecting the skin with approximately one million new cases annually, across more than 70 countries ^1^. In the Ethiopian highlands, up to 29 million people are at risk with an estimated 20,000-50,000 annual cases^2^, predominantly affecting young adults and children^3^. Facial lesions represent 45 – 97.6% of Ethiopian cases ^4^ and active lesions or disfiguring scars can lead to stigma, social exclusion, and poor mental health ^5^. Current treatments remain inadequate, with 32-38% failure rates ^6,7^ and treatment related morbidity that further compromises quality of life ^8^.

In Ethiopia, *L. aethiopica* is the dominant causative agent of CL producing an unusually broad range of clinical presentations ^9^, from self-healing localized lesions to chronic disfiguring disease. However, the immunological mechanisms underlying this remarkable heterogeneity remain unknown. CL due to *L. major*, *L. donovani* ^9,10^ and *L. tropica* ^11^ infections have also been reported (Hailu et al., 2006). Regardless of the infecting species, clinical cases are typically categorised using nomenclature from South American CL: localized (LCL), mucocutaneous (MCL) and diffuse (DCL) cutaneous leishmaniasis ^12^. LCL is most prevalent, but mucosal disease is also frequent^7,13^, while DCL is rare and presents as non-ulcerating nodules ^14–16^. Learning from studies on CL in Latin America, protective immunity is believed to require Th1-polarized CD4^+^ T cell responses and nitric oxide and reactive oxygen species-producing macrophages ^17^ while immunosuppressive cytokines (IL-10, TGF-β) ^12,18,19^ and “anergy” are associated with progressive DCL^20^. However, whether the South American terminology and associated immunopathological paradigms apply to Ethiopian CL has recently been questioned ^21^. Understanding whether the different clinical outcomes seen in Ethiopian CL reflect distinct immunopathological processes (as described in Latin America) or a spectrum of a single disease mechanism is critical for developing targeted immune-based therapies.

Despite being first reported in the 1930s ^22,23^, the immunopathogenesis of *L. aethiopica* disease remains poorly characterised at the tissue level. Histologically, Ethiopian CL lesions show predominantly dermal lymphohistiocytic inflammation with variable presence of granulomas^24^, neutrophil-mediated tissue damage via death ligands (FasL, TRAIL)^25^ and elevated arginase in lesions^26^. A recent analysis of plasma chemokines and cytokines and parasite sequencing failed to identify systemic immune signatures or parasite genetic factors associated with different clinical presentations^27^ suggesting that pathological mechanisms may be localized to lesional tissue. Yet the spatial organization of immune cells, stromal and epithelial compartments, and mechanisms driving tissue destruction versus healing responses remain uncharacterized.

In this study, we used spatial transcriptomics to comprehensively characterize the immunopathological landscape of CL lesions from five Ethiopian patients. We identified five fundamentally distinct pathotypes characterized as: organized cytotoxic responses with tertiary lymphoid structures, fibrotic wound healing, granulomatous inflammation with pro-inflammatory or M2 polarised myeloid cell inflammation, and epithelial cell hyperplasia. By defining the spatial organization of these responses and the molecular programs within distinct tissue microenvironments, our findings provide a framework for understanding disease heterogeneity and a first step towards immune-based classification of Ethiopian CL.

## Results

### Distinct transcriptomes in lesional and non-lesional skin of CL patients

To define the cellular landscape of the lesional microenvironment associated with Ethiopian CL, we recruited five male patients (P1-P5; median age 21 years, inter-quartile range [IQR]: 20-23) from diverse occupational and educational backgrounds from Gondar city in the Amhara Province, northern Ethiopia (**Figure 1A; Supplementary Table 1**). A schema indicating the methods and workflow of the study is provided in **Figure 1B**. Lesion appearance (**Figure 1C**) was documented along with detailed clinical and laboratory findings for each patient at baseline. Common lesion characteristics included plaque formation (3/5 of all patients), scaling (4/5), crusting (5/5), swelling (5/5), and erythema (4/5), with some cases showing additional features such as hyperpigmentation (1/5) and signs of bacterial infection (2/5). *Leishmania* species were confirmed in three patients: two *L. aethiopica* (P1, P2) and one *L. tropica* (P5) (**Table 1**).

**Figure 1.**
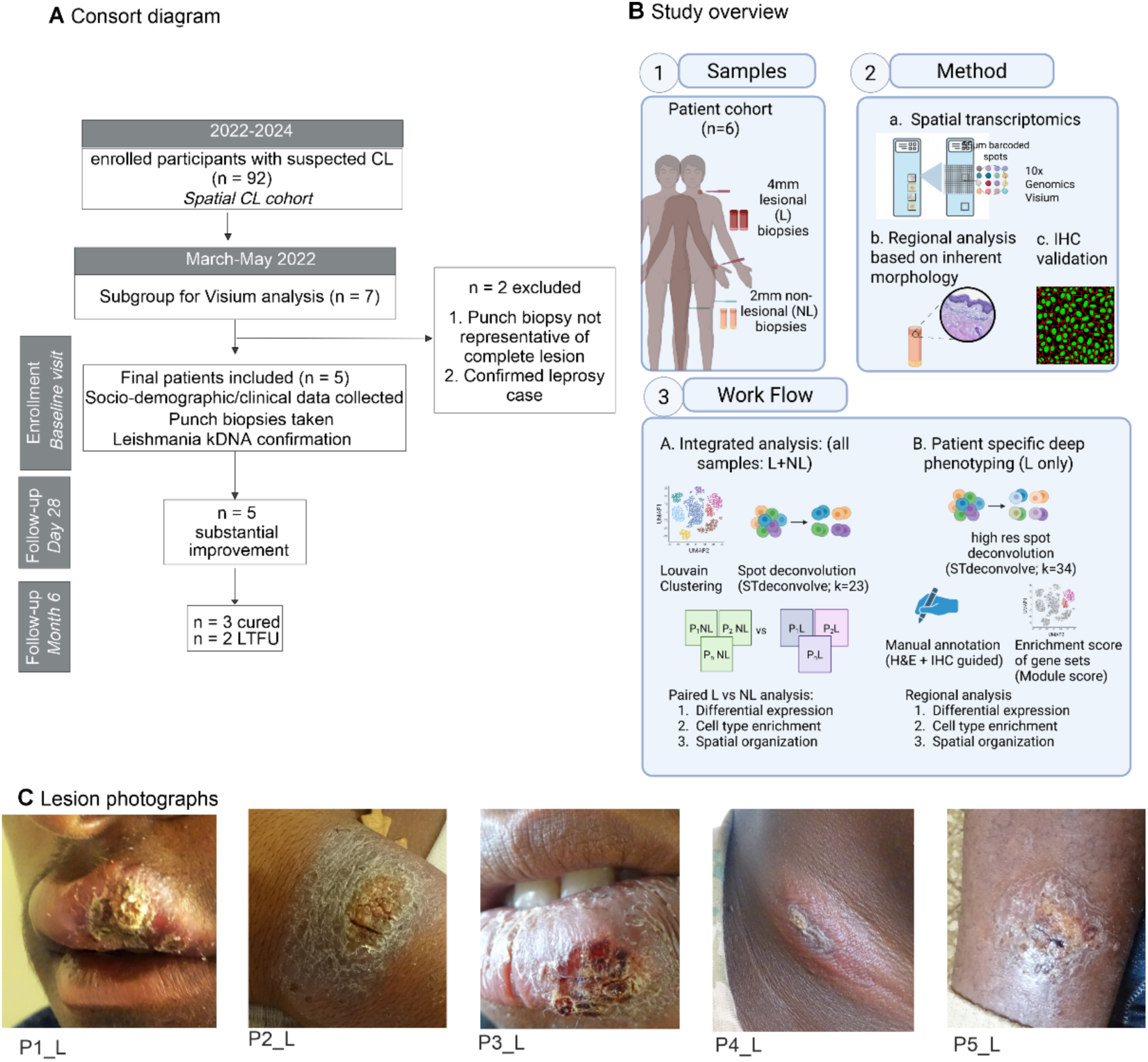
Study design and patient cohort characteristics. **A**, CONSORT diagram showing patient recruitment, inclusion criteria, and final study cohort. **B,** Schematic representation of study design showing patient cohort, types of biopsies collected, and analysis workflow of spatial transcriptomics data. **C,** Lesion photographs of the 5 patients included in the study.

**Table 1.** Clinical and laboratory data at baseline visit. Clinical characteristics of patients enrolled in the study.

| PID* | Duration of lesion in months | Number of active lesions | Lesion size index lesion (cm2) | Location of index lesion | Clinical classification | Characterization of index lesion | Location of other lesions | Characterization of other lesions | Scars | other dermatological conditions | Parasitemia microscopy grade | kDNA PCR# (Ct) | Parasite species typing<sup>Δ</sup> |
| --- | --- | --- | --- | --- | --- | --- | --- | --- | --- | --- | --- | --- | --- |
| P1 | 6 | 1 | 4 | lip | MCL | crusted, macrocheilial, swollen, erythematous, | NA | NA | No | acne vulgaris | 4+ | 9.94 | *L. aethiopica* |
| P2 | 26 | 1 | 15 | arms | LCL | plaque, scaly, crusted, swollen, erythematous, hyperpigmented | NA | NA | No |  | 0 | 27.31 | *L. aethiopica* |
| P3 | 4 | 3 | 8 | lip | LCL/MCL | plaque, scaly, crusted, dry, macrocheilial, swollen, exfoliative, | arms, ear | nodular, scaly, crusted, dry, swollen, erythematous | No |  | 0 | 33.42 | Not done |
| P4 | 11 | 1 | 21 | neck | LCL | nodular, scaly, crusted, dry, swollen, erythematous | NA | NA | No | acne vulgaris | 0 | 29.03 | Not done |
| P5 | 1 | 3 | 20 | legs | LCL | plaque, scaly, crusted, dry, swollen, super-infected | ear, chest | papular, plaque, erythematous | No |  | 0 | 20.03 | *L. tropica* |

\*All patients had CL as primary infection with no previous history of CL lesions; PID: Patient identifier; Ct (cycle threshold) value; #kDNA PCR = kinetoplast DNA polymerase chain reaction Δverified either with SureSelect or high resolution melt PCR or ITS-1 amplicon seq

To compare the spatial organisation of immune and stromal cells between lesional and non-lesional skin, we obtained one lesional and one non-lesional biopsy at baseline from each patient and performed spatial transcriptomics on the 10x Genomics Visium platform (**Figure 2**). After quality control, a total of 5562 spots were retained and pooled for downstream analyses. The total number of genes per spot was consistently higher in lesional compared to non-lesional skin, reflecting the increased cellularity of lesional samples (**Supplementary Figure 1A**).

**Figure 2.**
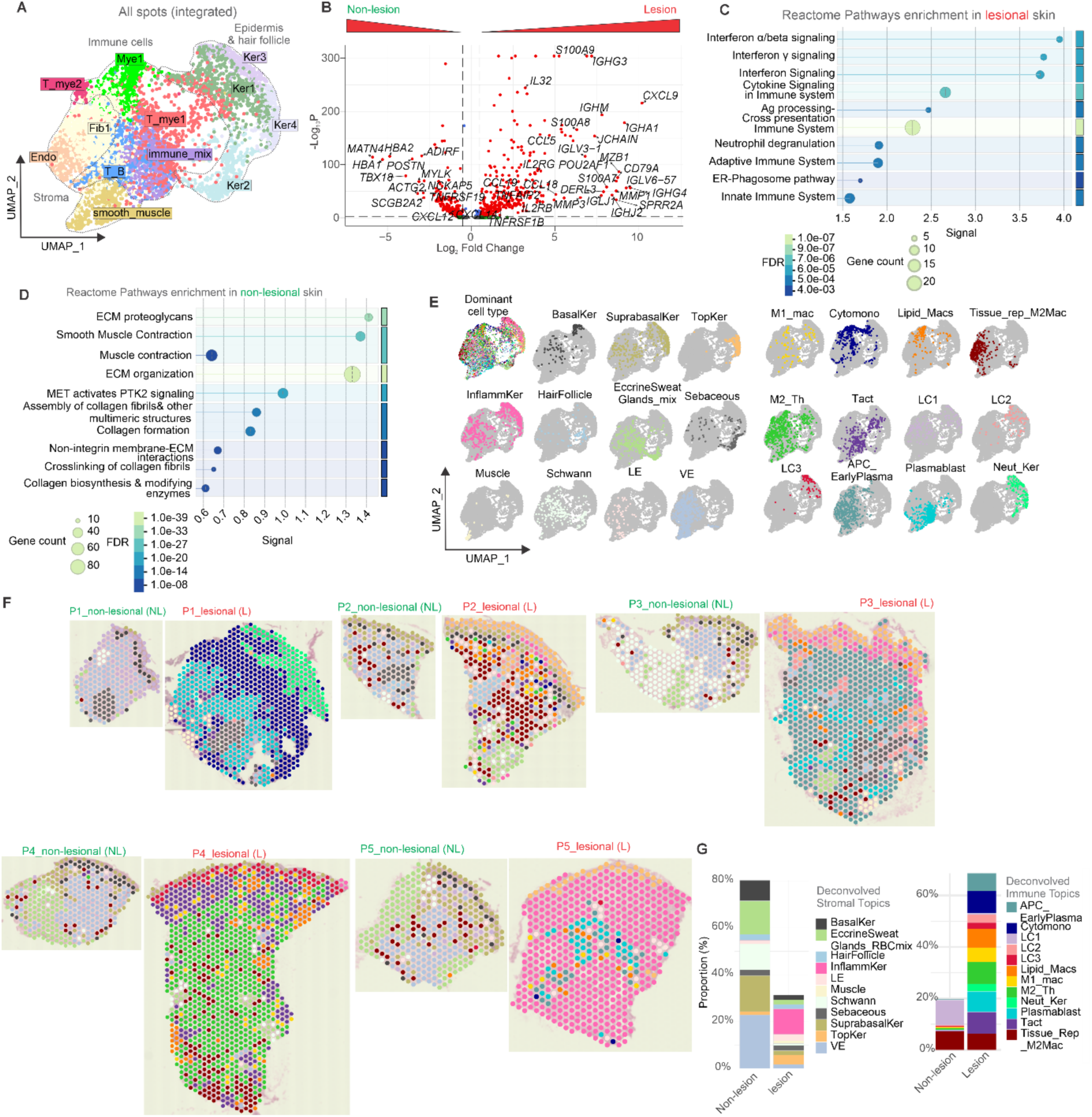
Global cellular heterogeneity and spatial organization in lesional vs non-lesional skin. **A**, Uniform manifold approximation and projection (UMAP) of pooled non-lesional and lesional skin spots labelled by major skin domains inferred from gene expression. **B**, Volcano plot of differentially expressed genes (DEGs) between non-lesional and lesional samples (red dots = adjusted P < 0.05, log2Fold > ±0.25, –log10p > 10). **C-D**, Top 10 Reactome pathways enriched in lesional (**C**) and non-lesional (**D**) skin. Bubble size = gene count; bars = signal strength (weighted harmonic mean of observed/expected ratio and – log(FDR)); pathways grouped by similarity (Jaccard index ≥ 0.8, ranked by p-value).**E**, UMAP coloured by dominant deconvoluted topic (highest proportion per spot) detected in each spot. **F**, Spatial maps of non-lesional and lesional skin (P1-P5) coloured by dominant topic per spot. **G**, Relative proportions of stromal (left) and immune (right) topic in non-lesional and lesional skin. All panels show pooled data from n=12 sections (6 non-lesional, 6 lesional from 5 patients) except (**F**), which shows individual patient maps.

Unsupervised clustering of all samples using the Louvain algorithm ^28^ identified 14 distinct clusters based on gene expression (**Figure 2A and Supplementary Figure 1, B**). Using cluster-specific gene expression signatures, we annotated three major spatial domains: i) epidermal and hair follicle, characterized by keratinocyte markers (in Ker1-4 clusters), ii) dermal immune enriched (immune_mix, T_B, T_mye1, T_mye2, Mye1 clusters) and iii) connective stromal tissue, rich in fibroblasts (fib1), smooth muscle cells (smooth_muscle cluster) and endothelial cells (Endo cluster). These annotations clearly separated in the UMAP visualization and spatial diagrams (**Supplementary Figure 1, C-D**) despite the integrated analysis of non-lesional (NL) and lesional (L) samples. This suggests a preservation of core transcriptional programs defining major skin compartments even in the disease state.

We next conducted differential gene expression analysis between all lesional and non-lesional skin to identify differentially abundant transcripts (**Figure 2B**). Lesional tissue showed marked upregulation of class switched immunoglobulin genes (*IGHA1*, *IGHG3*, *IGHG4*, *JCHAIN*), reorganisation of extracellular matrix (ECM; *MMP3*, *MMP1*), antimicrobial response (*SPRR2A*) and inflammatory markers (*S100A7*, *S100A8*, *CCL5*, *CXCL9*, *IL32*, *CCL18*, *CCL19*), while non-lesional skin was enriched for structural and stromal genes including *POSTN*, *MATN4*, *ADIRF* and *TBX18*. Gene set enrichment analysis (GSEA) revealed significant Reactome Pathway enrichment in lesional skin associated with interferon and other cytokine immune responses, antigen processing and presentation, neutrophil degranulation and ER (Endoplasmic Reticulum)-phagosome pathway (**Figure 2C**). In contrast, GSEA of non-lesional skin reflected the dominant stromal cell composition (**Figure 2D**).

Visium spots (55µm diameter) represents a tissue niche where 5-12 cells are co-localised. To estimate which cell types were contributing to the changes in transcriptomes of lesional and non-lesional tissue, we used a reference-free approach (STdeconvolve) based on inherent gene expression profiles to identify the cell-type composition of each spot^29^. Based on chosen resolution (see methods) STdeconvolve is able to generate gene expression profiles (termed “topics”) that are either lineage-enriched or mixed i.e. they contain genes associated with multiple lineages. Mixed topics thus represent either spatial co-occurrence of cell types or the presence of shared transcriptional programs active in multiple lineages. Using this deconvolution approach, we estimated cell type proportions for each spot. At a coarse resolution (K=23), we identified 11 stromal / epidermal and 12 immune cell topics, (see gene expression of topics shown in **Supplementary Figure 1, E-F** for terminology and gene profile defining each topic in **Supplementary Table 2**). For visualization purposes, we assigned each spot to its dominant cell type (defined as the cell type with the highest estimated proportion per spot; **Figure 2E-F**). Spatial mapping of dominant topics per spot demonstrated relatively uniform composition across non-lesional samples (evidencing efficient batch-correction), in contrast to the marked heterogeneity observed across lesional samples, with each patient displaying distinct spatial patterns (**Figure 2F**). Aggregating the estimated cell type proportions by immune versus stromal/epidermal topics revealed that non-lesional skin was predominantly stromal/epidermal, while lesional skin was largely immune (**Figure 2G)**.

To characterize topic composition differences between paired non-lesional and lesional skin samples, we performed linear mixed effects modelling ^30^ on spot-level deconvolution data (n=5,562 spots), accounting for patient-specific effects. This revealed striking differences in topic abundance (20/23 topics, p_adj < 0.05; log2FC: –4.06 to +6.62). Lesional and non-lesional tissues showed expected enrichment for immune and inflammatory topics and structural and epithelial topics, respectively (**Figure 2G** and **Supplementary Table 3**). These findings demonstrate that while the lesional tissue is highly heterogenous across different patients, each is characterized by increased immune cell infiltration when compared to the paired non-lesional tissue.

### Inter-patient variation in lesional architecture

To dissect the heterogeneity of each lesion in greater detail, we subclustered only the lesional tissues (**Supplementary Fig 2A**) and used STdeconvolve (resolution K=34) to obtain lesion specific topics. We identified 28 lineage-specific topics and six mixed topics with signatures from multiple cell types (**Figure 3, A-B, Supplementary Fig 2B-C and Supplementary Table 2**).

**Figure 3.**
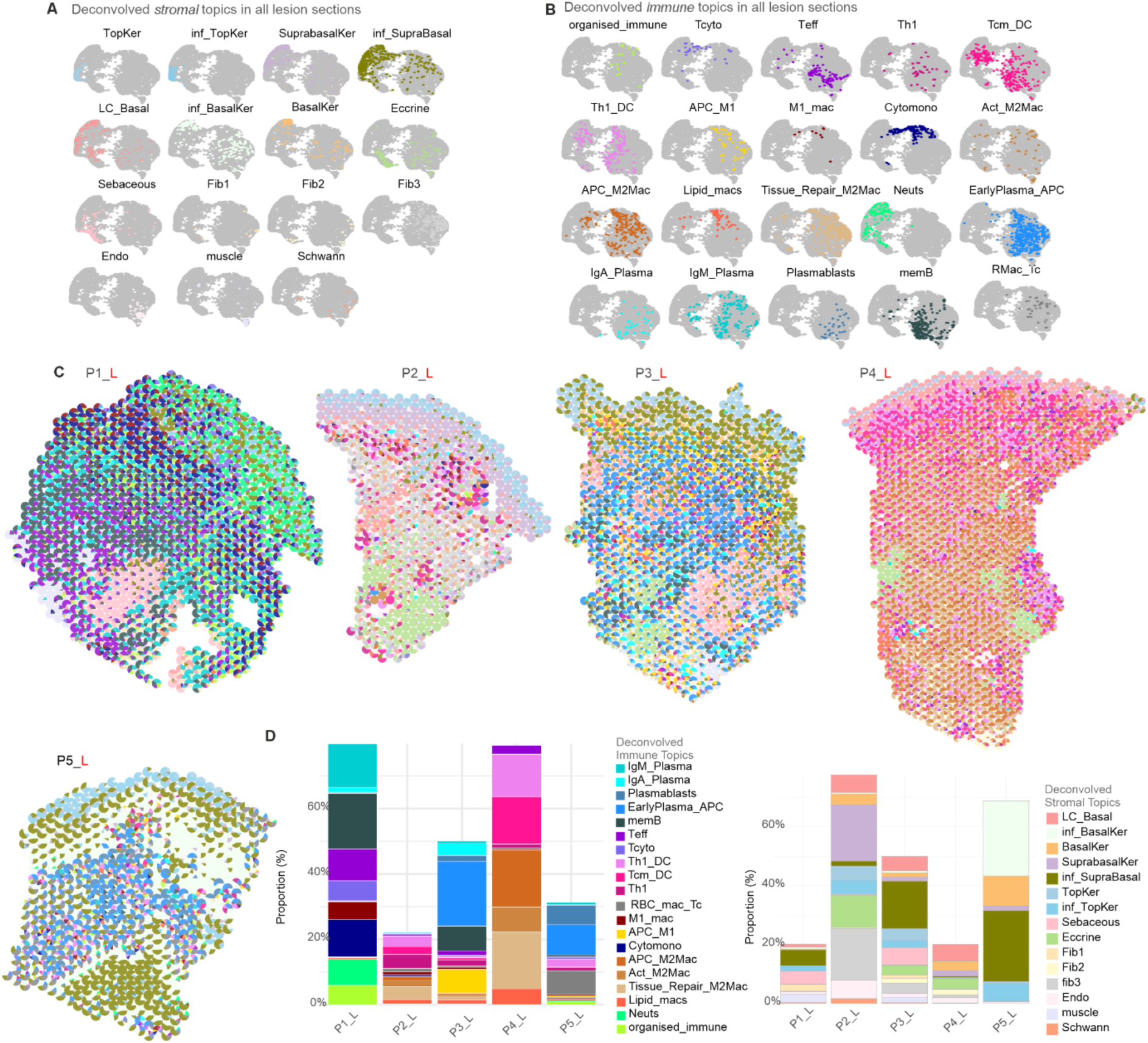
Cell type deconvolution reveals patient-specific patterns of immune and stromal composition in lesional tissue. **A-B**, Separate UMAPs for each stromal (**C**) and immune (**D**) topic, with spots colored by topic dominance. **C,** Spatial maps of each lesion with pie charts showing topic composition per spot. **D**, Relative proportions of immune (left panel) and stromal (right panel) topics per lesion.

Topic proportions per spot revealed distinct immunological features across patients (**Figure 3C-D**). P1 exhibited enrichment of B/plasma cells, diverse T cell subsets, proinflammatory macrophages, and neutrophils, suggesting a proinflammatory response. P2 showed high stromal topic contribution with Th1 predominating among immune topics. P3 was characterized by antigen-presenting cells while P4 had predominantly alternatively activated macrophages with T cell-DC mixed topics. P5 displayed the highest proportions of plasma cell, plasmablast, Cytomono, and inflammatory keratinocyte topics. To further characterize these differing immune microenvironments, we performed detailed analyses focusing on the dominant pathological features of each patient (**Figures 4-8**).

**Figure 4.**
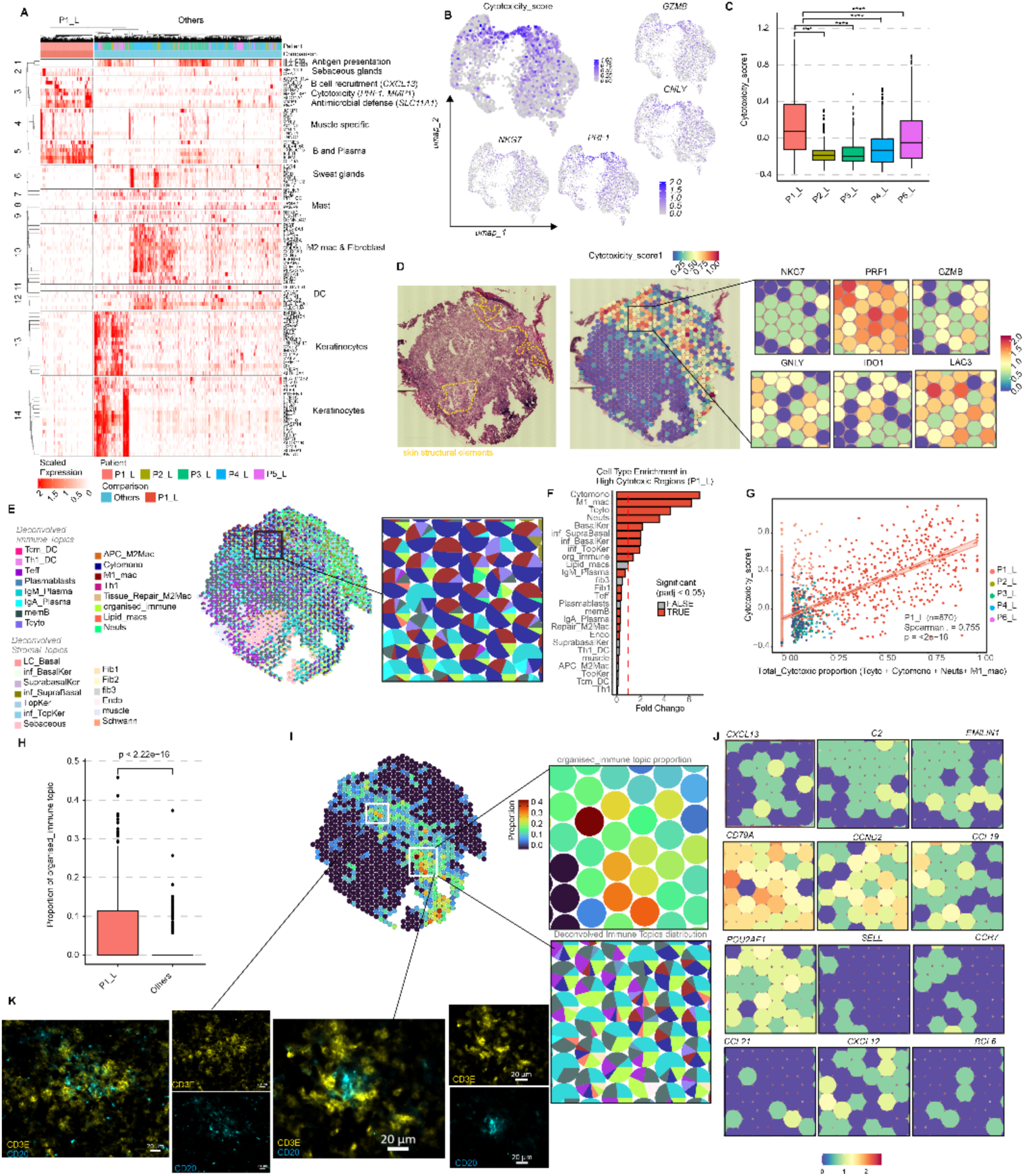
Cytotoxic immune response and tertiary lymphoid structures in P1. **A**, Heatmap of top 100 DEGs in P1_L vs other lesional skin (P2-P5 L) (padj < 0.05, log2FC > ±0.25; Wilcoxon) **B**, UMAP of cytotoxicity signature in lesional skin (P1-P5). Top: composite score; right/below: four cytotoxic effector genes. **C**, Cytotoxicity score across all patients (Wilcoxon; P1_L vs P2-P5 L). **D**, Spatial map of cytotoxicity score in P1_L (middle), underlying H&E histology (left panel) and zoomed view of a representative area showing individual gene expression for selected genes (right). **E,** Cell type deconvolution (pie charts) of representative area (from **D**). **F**, Fold enrichment of topics in high (≥Q3, top 25%) vs low (<Q3, bottom 75%) cytotoxic regions of P1_L (Wilcoxon with Bejamini Hochberg correction; red dotted line: log2FC =1). **G**, Correlation between total cytotoxic cell abundance (Tcyto, Cytomono, M1_mac, Neuts) and cytotoxicity score. Points coloured by patient; P1_L (95% CI); ρ = Spearman’s rank. **H**, Organised_immune topic proportion in P1 L vs P2-P5 L (Wilcoxon). **I**, Organized immune topic distribution in P1_L across full tissue (left), magnified region (top right), and other topics as pie charts (bottom). **J**, tertiary lymphoid structures (TLS) hallmark gene expression in representative area from (**I)**. **K**, Multiplex immunofluorescence (IF)of two putative TLS from P1_L showing CD3E+ T cells (top) and CD20+ B cells (bottom) in magnified regions. Scale bar, 20µm. **p < 0.01, ****p < 0.0001

### Cytotoxic T cell responses and lymphoid neogenesis

Patient P1 was enriched for inflammatory cells populations in lesional tissue (**Supplementary 3A**) and differential expression analysis against all other patients revealed elevated expression of the cytotoxic effector (*PRF1*), B cell and plasma cell-associated genes (*IGLV4-60*, *TXNDC15*, *IGLJ1*, *IGHM*, *CD79A*), B cell-attracting chemokine *CXCL13*, enrichment of antimicrobial response genes (*HS3ST3B1*, *SLC11A1*), and ECM remodelling enzymes (*MMP1*) (**Figure 4A**).

We first calculated a cytotoxicity score using the average expression of established cytotoxic markers (*GZMB*, *GNLY*, *NKG7*, *PRF1*) to compare cytotoxic activity across all patient lesions (**Figure 4B** and **Supplemental Figure 3B**). P1 exhibited a higher cytotoxicity score compared to all other patients (**Figure 4C**). Spatial mapping revealed that high-cytotoxicity areas (defined as >75th percentile of cytotoxicity score) spanned the papillary dermis and extended into the reticular dermis. These regions showed significant enrichment of interferon-responsive genes (*ISG15*, *RSAD2*, *CXCL11*), antimicrobial factors (*CTSL*, *SLC11A1*), and immune check point markers (*IDO1*, *LAG3*) (**Figure 4D** and **Supplemental Figure 3C**). Deconvolution of P1-specific high cytotoxicity spots revealed enrichment (p < 0.001, Wilcoxon) of lineage-enriched topics: Cytomono (6.93-fold), Tcyto (4.48-fold), M1_mac (6.13-fold), and Neuts (3.53-fold) (**Figure 4E-F**, **Supplemental Figure 3D and Supplementary Table 4)**. The combined proportions of these four topics correlated significantly with cytotoxicity scores in P1 (**Figure 4G**) but not in other patients, indicating that this specific cellular composition is uniquely associated with P1.

The second major feature associated with P1 was the presence of a mixed topic (organised_immune) with co-occurrence of genes associated with B cells, T cells and APC. This topic is indicative of tertiary lymphoid structures (TLS), ectopic lymphoid aggregates that form at sites of chronic inflammation. Among all patients, P1 exhibited the highest proportion of the organized_immune topic (p < 0.001, Wilcoxon) (**Figure 4H**), with spatial distribution concentrated in the reticular dermis (**Figure 4I**). To characterize these microenvironments further, we mapped canonical TLS markers, revealing coordinated expression of *CXCL13* and *CD79A* (B cell recruitment), *POU2AF1* (a transcriptional co-activator required for germinal centre formation), C2 (complement-mediated antigen presentation), *CCND2* (cell cycle activity), lymphocyte recruitment chemokines and receptors (*SELL*, *CCL19*, *CCR7*, *CXCL12*), extracellular matrix (Fibroblastic reticular cell marker, *CCL21* and *EMILIN1*) and *BCL6* (a marker of mature germinal centres) (**Figure 4J**). The coordinated spatial expression of these markers confirms the presence of organized TLS in P1. To confirm at the protein level, immunohistochemistry revealed aggregated CD20⁺ B cells forming a central follicular core surrounded by CD3⁺ T cells, indicative of organised TLS and recapitulating the characteristic zonation of organized lymphoid structures ^31,32^ (**Figure 4K**). Notably, P1 had a higher parasite load than others (microscopy grade 4+, kDNA PCR ct= 9.94; **Table 1** and **Supplementary Figure 3E**). In summary, P1 was characterized by high skin parasite load, a robust cytotoxic immune response, and the presence of TLS.

### Fibrotic remodelling with preserved stromal architecture

Comparison of P2 to all other lesional samples revealed upregulation of collagen genes (*COL6A1*, *COL6A2*, *COL1A1*, *COL3A1*, *COL1A2*), fibrotic markers (*PDGFRB*), ECM components (*FBLN2*, *FBLN1*, *DCN*, *SPARC*), remodelling enzymes (*MMP2*, *TIMP2*, *TIMP3*), and the myofibroblast marker *ACTA2*, collectively indicating active wound healing and fibrotic remodelling (**Figure 5A)**. These findings corroborate the clinical and histological presentation of this patient, including a prolonged lesion duration of 26 months with hyperpigmentation and evident fibrosis (**Figure 1D and Figure 5B**).

**Figure 5.**
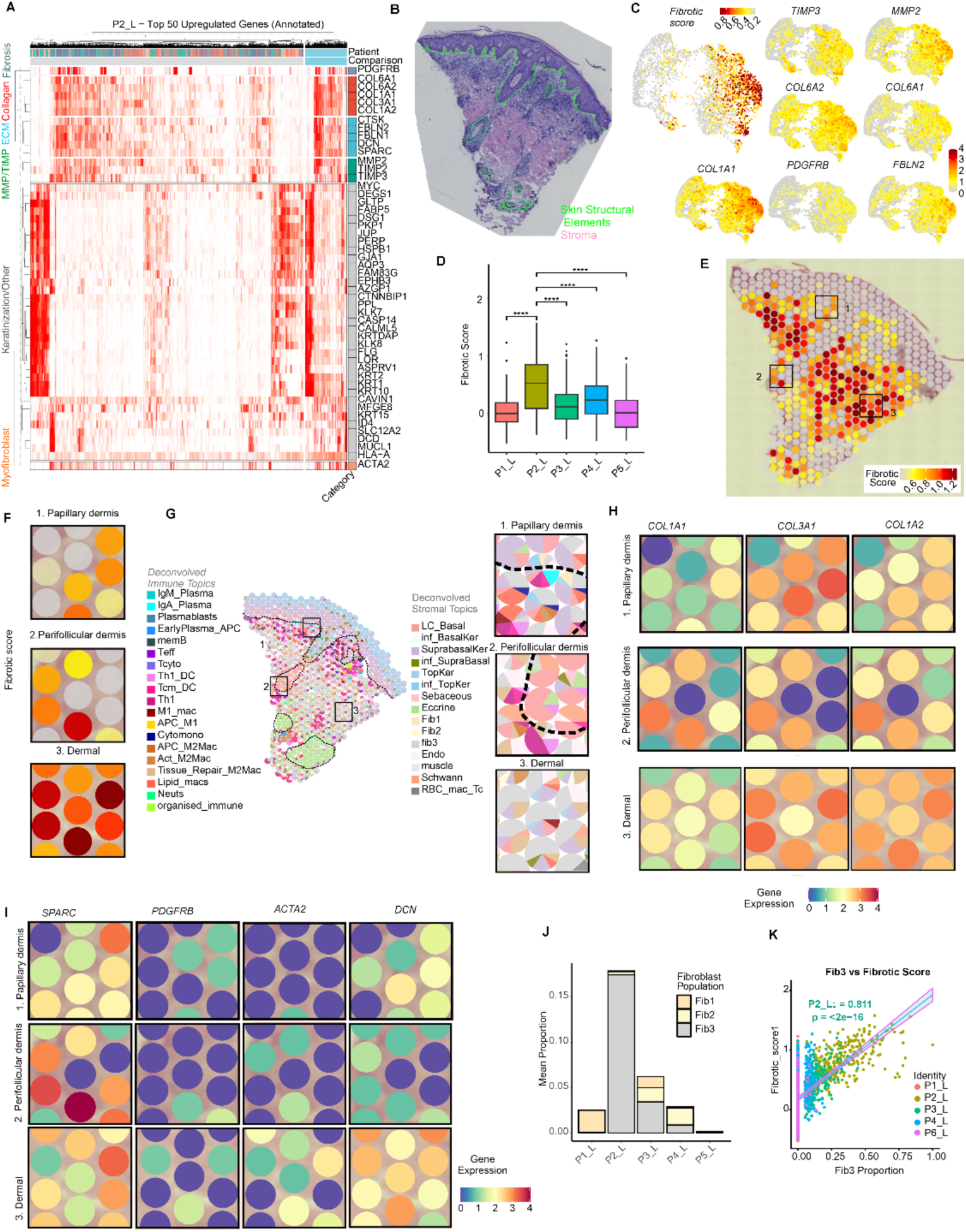
P2 shows extensive fibrotic remodeling with reduced immune infiltration. **A**, Heatmap of top 50 upregulated genes between patient P2_L vs other lesional skin (P1, P3-P5 L) with functional annotation (padj < 0.05, log2FC > ±0.25; Wilcoxon). **B**, H&E histology from P2_L. green dotted line: skin structural elements: **C**, UMAP of fibrosis signature (in lesional skin (P1-P5). Top: composite fibrosis score; right/ below: 7 selected individual fibrotic genes. Colour scale: 25th-95th percentile). **D**, Fibrotic score comparison across all lesional skin (Wilcoxon; P2L vs others). **E**, Spatial map of fibrotic score in P2_L (colour scale: q25-q95 for P2 only) with three representative regions (black boxes): 1-papillary dermis, 2-perifollicular dermis, 3-dermal region. **F**, Enlarged views of regions 1-3 showing compartment-specific patterns. **G**, Cell type deconvolution (pie charts) on regions from (**F**). **H-I**, Spatial expression of selected fibrotic genes in regions from (**F**). **J**, Mean proportion of Fib1, Fib2 and Fib3 fibroblast deconvoluted topics in lesional skin (P1-P5). **K,** Correlation between Fib3 proportion and fibrotic score across all spots (P1-P5). Points coloured by patient; trendline: P2_L (95% CI); ρ = Spearman’s rank).

To validate this observation, we created a fibrotic score based on average expression of *TIMP3*, *MMP2*, *COL6A2*, *COL6A1*, *COL1A1*, *PDGFRB*, *FBLN2* genes and compared scores across different patients. P2 had the highest fibrotic score (**Figure 5C-D** and **Supplemental Figure 4**). Spatial plotting of the fibrotic score revealed extensive fibrosis in the reticular dermis consistent with stromal eosin staining in histology (**Figure 5B, E**). To look at spatial gene expression of fibrotic genes and whether they differed across different sites we then concentrated on three areas in the dermis, 1, papillary dermis, 2, perifollicular dermis and 3, reticular dermis (**Figure 5F**). The papillary and perifollicular dermis were also associated with immune infiltrate (**Figure 5G**) and therefore the reticular dermal spots which had the lowest immune composition had the highest fibrotic score. While most of the fibroblast genes were expressed in all three areas (**Figure 5H-I**), transcripts for the myofibroblast gene *ACTA2* and fibrosis gene *PDGFRB* were of lowest abundance in the papillary dermis and perifollicular dermis, respectively.

Among the three fibroblast-enriched topics (Fib1-3), P2 showed enrichment of Fib3, which expressed ECM remodelling genes (*ELN*, *PODN*, *MFAP4*, *THBS2*), matrix-degrading enzymes (*MMP2*, *CTSK*), and myofibroblast markers (*SFRP2*, *LRRC15*) (**Figure 5J**). The proportion of Fib3 strongly correlated with fibrotic score (Spearman’s ρ = 0.8, p < 0.001), suggesting this population is associated with fibrotic remodelling (**Figure 5K**). Together, these findings demonstrate that P2 is characterized by extensive fibrotic remodelling and reduced immune infiltration, as might characterize a wound healing phenotype.

### Granulomatous inflammation with proinflammatory characteristics

Histological examination of P3 revealed extensive immune infiltration with multiple discrete granulomas throughout the dermis. Granulomas were characterised by an epithelioid CD68^+^ macrophage-rich core and a lymphocytic cuff, with abundant CD8A^+^ cells (**Figure 6A and B)**. To dissect the molecular and cellular features of these granulomas further, we manually annotated each granuloma in the spatial transcriptomics data using the underlying tissue morphology (**Figure 6C**). Spatial maps of the proportion of deconvoluted topics in spots representing two representative granulomas revealed enrichment of macrophage, B cell and T cell topics, with spatial heterogeneity in distribution within each granuloma (**Figure 6D**). Spatial gene expression mapping showed coordinated expression of markers for foamy macrophages (*CHI3L1*, *LIPA*, *APOE*), MHC class II molecule (*HLA-DQA1)*, antimicrobial genes (*MPEG1*, *SOD2*), and chemokines involved in T cell recruitment and activation (*CCL5*, *CXCL9*, *LTB*) (**Figure 6E**). These genes were significantly enriched in granulomatous areas compared to non-granulomatous regions (**Figure 6F**). Cell type enrichment analysis revealed that granulomas were predominantly composed of lipid-associated / tissue adapted macrophages (*CHIT1*, *CHI3L1*, *LIPA*, *CYP27A1*, *CD68*; 12.8-fold, p < 0.001), cytotoxic monocytes (*APOE*, *PRF1*, *LYZ*; 11.5-fold, p < 0.001), proinflammatory M1 macrophages (*TCN2*, *G0S2*, *CD68*; 5.0-fold, p < 0.001), and effector T cells (*FCMR*, *CXCR3*, *LTB*, *CCL5*; 3.0-fold, p < 0.001) (**Figure 6G**).

**Figure 6.**
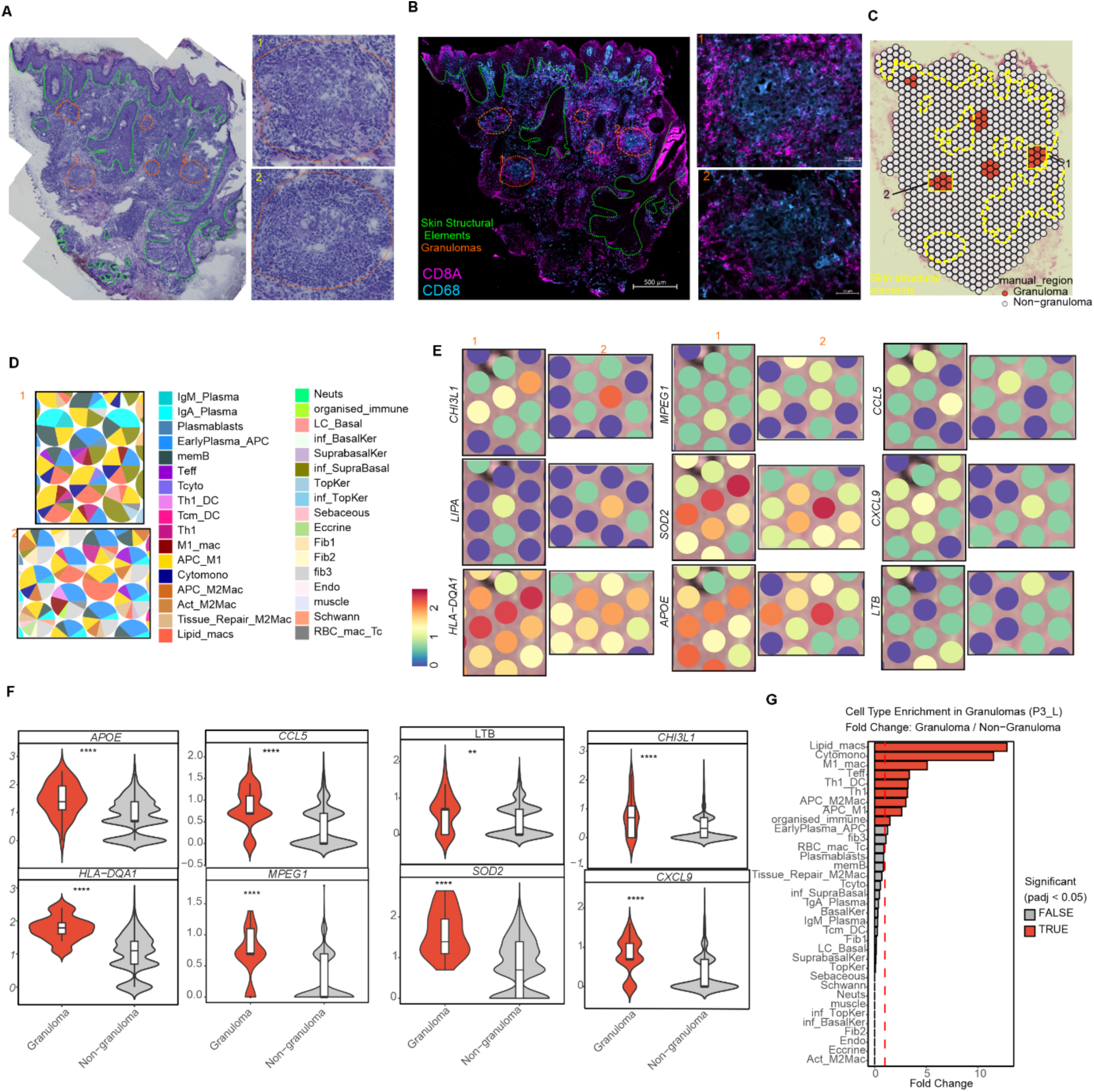
Patient P3 L exhibits distinct granulomatous inflammation with enriched antigen presentation machinery and lipid-associated macrophages. **A**, H&E staining of P3_L tissue. Right: magnified views of representative granulomas (regions 1-2). **B**, Multiplex IF staining of CD68 (myeloid; blue) and CD8 (T cells; magenta) of P3_L. Right: high-magnification views of regions 1-2 from (**A**).**C,** Spatial map of P3_L showing granulomatous (red) and non-granulomatous (gray) regions based on underlying histology. Yellow dotted lines and black boxes: structural elements and regions 1-2 (from **A**). **D,** Cell type deconvolution (pie charts) for regions from (C) **E**, Spatial expression of selected marker genes in regions 1 and 2 from (**C)**. **F**, Expression of genes from (**E**) in granulomatous (red) vs non-granulomatous (gray) regions (Wilcoxon). **G**, Fold change in cell type proportions between granulomatous and non-granulomatous regions in P3_L. Red: significant enrichment (padj < 0.05, Wilcoxon); red dotted line drawn at log2FC =1.0. In panels (**C-D**) granulomatous regions marked with orange dotted lines; skin structural elements (epidermis, hair follicles) outlined in green. **p < 0.01, ****p < 0.0001

Additionally, differential gene expression analysis between P3 and all other patients identified a marked B cell infiltration in this patient with abundant transcripts for class-switched immunoglobulin genes (**Supplementary Figure 5, A-C**). Together, these findings demonstrate that P3 is characterized by the presence of organized granulomas with proinflammatory / remodelling characteristics, and that are embedded in a microenvironment with prominent B cell infiltration.

### Granulomatous inflammation with M2 myeloid cell polarization

Similar to P3, histology from P4 indicated a prominent granulomatous response (**Figure 7A**), but differential gene expression analysis revealed a strikingly different surrounding microenvironment (**Figure 7B** and **Supplemental Figure 6A**). All P4 spots had enrichment of lysosomal genes (*MARCO*, *CTSH*, *MPEG1*, *TMEM176B*, *CTSZ*), MHC-II and antigen-presenting machinery genes (*CD4*, *HLA-DMB*, *HLA-DPA1*, *IRF8)* chemokines and immune mediators (*CXCL9*, *TRAC*, *FYB1*, *C1QA*, *LCP1*, *FCER1G*, *CXCL14*), complement genes (*SERPINB1*, *PSGL2*, *C3*), myeloid markers (*CLEC10A*, *MARCO*, *MS4A6A*), lipid metabolism genes (*PLA2G2D*, *PLA2G2A*, *PLB7*, *SLC40A1*), and ECM remodelling factors (*TGFB1*, *TIMP1*, *TIMP3*, *FN1*, *CLEC3B*, *MFAP5*, *IGFBP6*), collectively indicating dominant infiltration by tissue-resident macrophage phenotypes with active tissue repair and remodelling programs. Conversely, P4 showed reduced expression of B cell and plasma cell markers (*IGHG1-4*, *POU2AF1*, *CD79A*, *DERL3*, *PRDM1*) and keratinocyte inflammatory genes (*KRT6B*, *S100A2/7/8*), suggesting minimal plasmacytic and epithelial inflammation compared to other patients (**Figure 7B**). These gene expression patterns suggested enrichment of alternatively activated myeloid phenotypes in P4.

**Figure 7.**
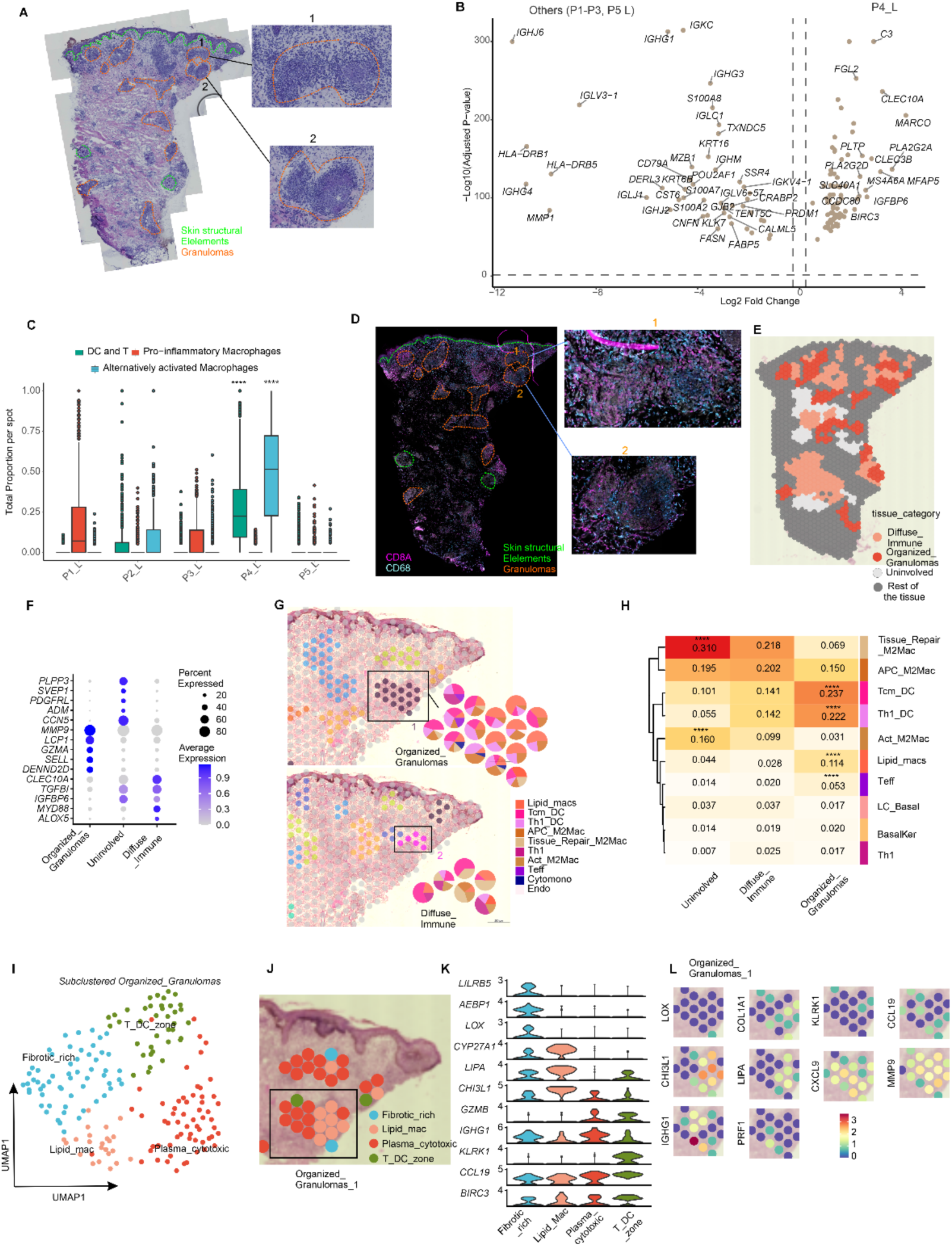
Patient P4_L demonstrates M2-polarized macrophage microenvironment with embedded granulomas. **A**, H&E of P4_L with magnified regions 1-2. **B**, DEGs between P4_L and other lesional samples (P1-3 & P5 L; top 50 genes labelled, Wilcoxon test). **C,** Comparison of macrophage, T and DC topic proportions across patients (pairwise Wilcoxon test). **D,** CD8A/CD68 multiplex IF on P4_L (right) with magnified regions 1-2 from (**A**) (left). **E,** Spatial annotation map categorizing immune infiltration patterns (organized granuloma, diffuse immune, uninvolved, other). **F,** Top 5 DEGs per annotation category. **G,** Representative organized immune (area 1) and diffuse immune (area 2) regions with deconvolved cell type pie charts showing distinct compositions. **H,** Mean cell type proportions across annotation categories (top 10 enriched types, pairwise Wilcoxon). **I-J,** UMAP subclustering of organized immune spots **(I**) and spatial projection onto area 1 (**J). K-L,** Top marker genes for subclusters (stacked violin plots, K) and their spatial expression in organized granuloma area 1 (**L**). (, ****p < 0.0001). In (**A**, **D**) Granuloma: orange dotted lines; skin structural elements (epidermis, hair follicles): green.

**Figure 8:**
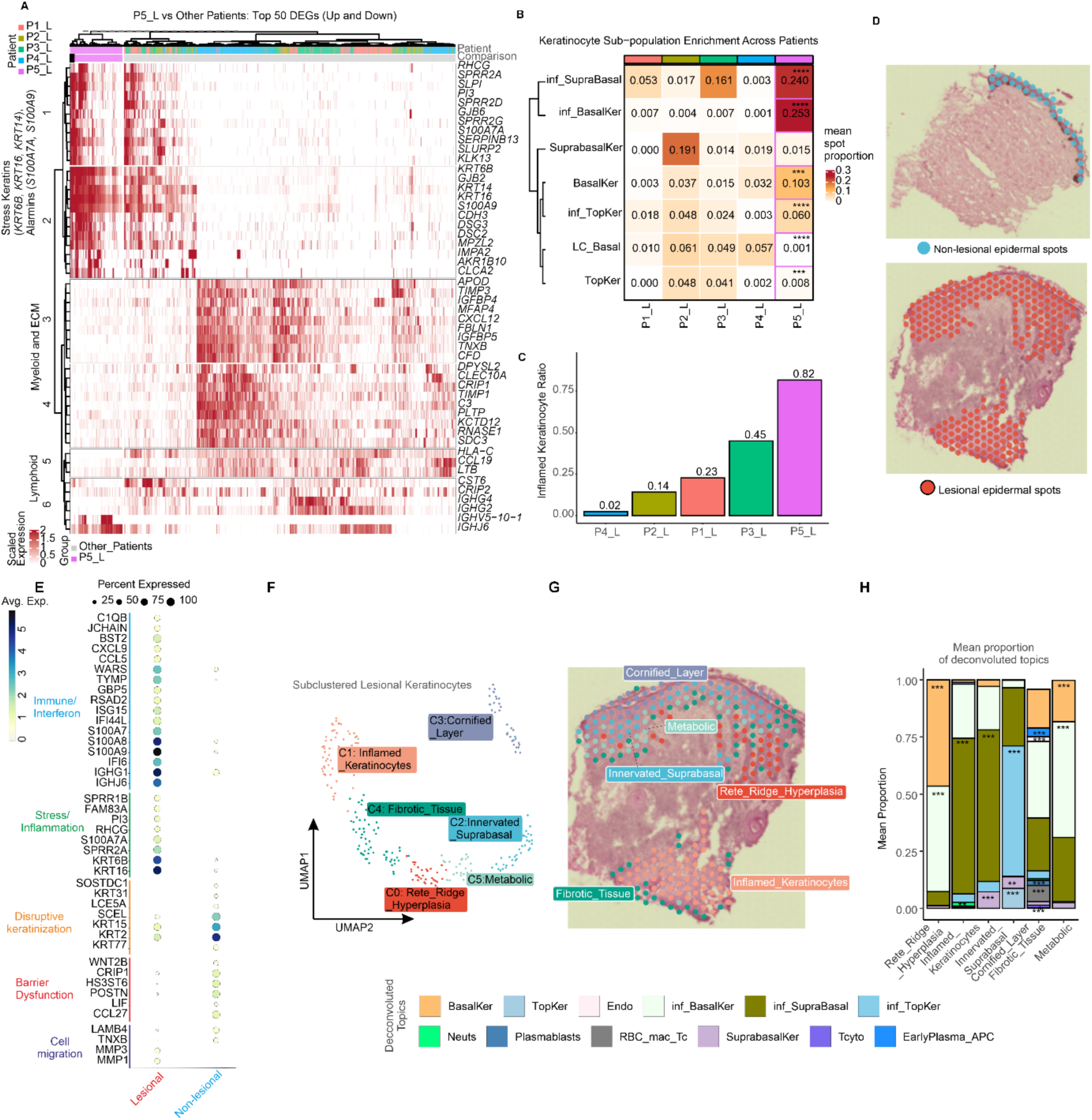
Spatial and molecular characterization of pseudoepitheliomatous hyperplasia in patient P5_L. **A**, Heatmap showing top 50 DEGs in P5_L vs other lesional skin (P1-P4_L); annotated by functional category. **B,** Heatmap of keratinocyte subpopulation enrichment across patients (mean proportions; pairwise Wilcoxon). **C,** Mean proportion of inflamed keratinocyte subtypes (inf_BasalKer, inf_SupraBasal, inf_TopKer) relative to total keratinocytes (inflamed and non-inflamed) per patient. **D**, Spatial map of P5 with H&E showing epidermal spots. Blue: P5_NL epidermis; orange/red: P5_L epidermis. **E** Marker gene expression between P5_L and P5_NL epidermal spots from (**D**), grouped by biological function. **F,** UMAP of P5_L epidermal spots showing clusters. **G,** Spatial distribution of the six epidermal subclusters from (**F**) overlaid on H&E**. H,** Mean proportions of deconvoluted topics (>1% abundance) per epidermal subcluster in (**F**). *p<0.05, **p<0.01, ***p<0.001.

To quantify macrophage phenotypes and T cell-dendritic cell interactions, we compared the combined proportions of macrophage and T-DC mixed topics across patients. P4 exhibited significantly higher proportions of topics enriched for alternatively activated macrophages (APC_M2Mac, Act_M2Mac, Tissue_repair_M2Mac, and Lipid_macs per spot) and the mixed Th1_DC and Tcm_DC topics compared to all other lesions (Wilcoxon rank-sum test, p < 0.001) (**Figure 7C**). In contrast, total proinflammatory macrophage topics (Cytomono, M1_mac, and APC_M1) were not significantly enriched in P4.

To determine whether this tissue microenvironment influenced granuloma form or function, we first identified granulomas in P4 by immunostaining, and annotated four distinct tissue compartments based on morphology: (1) organized granulomas with defined epitheliod macrophages and lymphocytic cuff (Organized_Granulomas, **Figure 7 D-E**), (2) diffuse peri-granuloma infiltrate without clear organization (Diffuse_Immune), (3) uninvolved stroma with minimal immune infiltration (Uninvolved), (4) remaining tissue (**Figure 7E**). These regions were associated with distinct molecular signatures reflecting lymphocyte recruitment, cytotoxicity and tissue remodelling in organized granulomas (*MMP9*, *LCP1*, *GZMA*, *SELL, DENND2D*) myeloid cell surveillance and regulation in diffuse immune areas (*CLEC10A*, *TGFB1*, *IGFBP6*, *MYD88*, *ALOX5*), and ECM genes in uninvolved stromal regions (*CCN5*, *ADM*, *PDGFRL*, *SVEP1*, *PLPP3*) (**Figure 7F**).

Mapping deconvoluted topics onto a representative granuloma and its surrounding diffuse immune region revealed higher proportions of lipid-associated macrophages, cytotoxic monocytes, effector T cells, and Th1-DC interactions within the organized granuloma compared to adjacent diffuse infiltrate (**Figure 7G**). To systematically characterize cellular enrichment patterns across all annotated regions, we performed cell type enrichment analysis comparing each region to organized granulomas. Tissue repair M2 macrophages (*MARCO*, *MS4A6A*, *CLEC10A*, *CD68*; 4.5-fold; padj<0.0001) and activated M2 macrophages (*CCL13*, *VSIG4*, *CD14*; 5.16-fold; padj<0.0001) were enriched in uninvolved stroma, while antigen-presenting M2 macrophages (*PLA2G2D*, *HLA-DQA1, CD68*) predominated in diffuse immune areas (1.34-fold; padj<0.001). In contrast, both Tcm-DC (*CCL19*, *IL7R*, *TRBC2*,

*CLEC10A; ∼*1.5 fold, padj<0.0001), Th1-DC (*CXCL9*, *IL4I1*, *CD4*, *CD3D; ∼*1.5 fold, padj<0.0001), and lipid-associated macrophages (4.12-fold, padj<0.0001) were most abundant in organized granulomas when compared to the neighbouring diffuse immune infiltrate (**Figure 7H** and **Supplemental Figure 6B**).

Finally, to identify potential spatial zonation within granulomas, we subclustered all granulomatous spots and examined their spatial distribution (**Figure 7I-J**). Based on gene expression profiles, we identified four distinct zones (**Figure 7I, Supplemental Figure 6C**): (1) a fibrotic zone characterized by ECM genes (*FN1*, *COL3A1*, *COL1A2*, *COL1A1*, *TIMP1*, *MMP2*); (2) a plasma cell-rich cytotoxic zone expressing immunoglobulins and cytotoxic effectors (*IGHG1*, *IGHA1*, *JCHAIN*, *GZMB*, *PRF1*); (3) a T cell-dendritic cell interaction zone marked by chemokines and T cell markers (*CCL19*, *CXCL9*, *GBP5*, *CCL17*, *CD3D*, *CD3E*); and (4) a lipid-associated macrophage zone with markers of foamy macrophages and tissue remodelling (*CHI3L1*, *LIPA*, *PLIN2*, *MMP9*, *WARS*, *APOE*, *CD68*). Spatial mapping of these subclusters onto a representative granuloma (organized_granuloma_1) revealed a structured architecture with distinct spatial compartmentalization (**Figure 7J**) and gene expression patterns (**Figure 7K-L).** The fibrotic zone showed highest expression of *LILRB5*, *AEBP1*, *COL1A1* and *LOX*. The lipid macrophage zone expressed *CYP27A1*, *LIPA*, and *CHI3L1*. The plasma-cytotoxic zone exhibited elevated *GZMB* and *IGHG1* and the T-DC zone showed enrichment of *KLRK1* and *CCL19*. Notably, *BIRC3*, an anti-apoptotic factor involved in NF-κB signalling, showed ubiquitous expression across all granuloma subclusters, indicating active cell survival programs throughout the organized structure. Spatial gene expression mapping confirmed these distinct molecular territories. Notably, we observed a broader *CXCL9* and *MMP9* expression indicating sustained immune activation and tissue remodelling throughout the granulomatous region (**Figure 7L**).

Together, these findings demonstrate that P4 exhibits a unique immunological architecture characterized by organized granulomas embedded within an M2-polarized tissue microenvironment. While P4 granulomas share structural features with P3, including epithelioid macrophages and lipid-associated macrophage signatures, differential expression analysis between P3 and P4 granulomas (**Supplemental Figure 6D**) revealed that P3 granulomas had more abundant transcripts for MHC class II genes (*HLA-DRB1*, *HLA-DRB5*) and immunoglobulins (*IGHG1*, *IGHG3*, *IGHG4*, *IGHJ6*, *IGHJ2*). P4 granulomas, in contrast, show higher complement activation (*C3*) and chemokine-mediated T cell recruitment (*CXCL9*, *CCL19*). Thus, P3 and P4 represent distinct forms or stages of granulomatous inflammation distinguishable by gene expression.

### Pseudo-epitheliomatous hyperplasia with marked immune infiltration and interferon response

The preceding analyses revealed distinct immune-centric pathologies across the four (P1-P4) patients. We next examined P5, the only patient with confirmed *L. tropica* infection. Comparative transcriptomic analysis of P5 against all other patient lesions revealed upregulated genes associated with keratinocyte hyperproliferation (*KRT6B*, *KRT14*, *KRT16*), antimicrobial defense and alarmins (*S100A7A/9, SLPI*, *PI3*, *SERPINB13*), cornification and cell adhesion and junction proteins (*DSG3*, *DSC2*, *CDH3*, *GJB2/6*) (**Figure 8A**) indicating epidermal hyperplasia as the dominant pathological feature of P5. Conversely, P5 showed reduced expression of ECM genes (*TIMP3*, *IGFBP4*/5 *MFAP4*, *FBLN1*, *TIMP1*), tissue-resident myeloid markers (*CLEC10A*, *C3*, *RNASE1*), immune genes (*HLA-C*, *CCL19*, *LTB*), and class-switched immunoglobulins (*IGHG2*, *IGHG4*).

Inflamed keratinocyte topics were strikingly enriched across all three epidermal layers in P5 (inf_BasalKer, BasalKer, inf_SupraBasal, inf_TopKer; padj<0.05, Wilcoxon; **Supplementary Table 5**; **Figure 8B**), while the LC_Basal topic was 50-fold reduced (padj<0.0001), suggesting Langerhans cell displacement. This pan-epidermal inflammatory signature indicates inflammatory hyperplasia rather than proliferative expansion Quantitatively, P5 exhibited the highest inflamed-to-total keratinocyte ratio among all patients (0.82; **Figure 8C**)

To characterize inflamed keratinocyte signatures, we manually annotated epidermal regions based on tissue morphology and extracted gene expression profiles from matched lesional (P5_L) and non-lesional (P5_NL) epidermal spots (**Figure 8D**). Differential expression analysis between P5_L and P5_NL epidermis revealed disruption of five major biological programs (**Figure 8E** and **Supplemental Table 6**): (a) immune/interferon infiltration, with increased expression of interferon-stimulated genes, immunoglobulins, immune mediators, and DAMPs; (b) stress and inflammation, with elevated stress keratins, cornified envelope precursors, and antimicrobial peptides; (c) disrupted differentiation, with reduced terminal differentiation markers; (d) barrier dysfunction, with loss of homeostatic signaling molecules including CCL27 and LIF, and structural genes; and (e) altered cell migration with upregulation of matrix remodelling genes.

To identify functionally disrupted keratinocyte compartments, we subclustered all epidermal spots from P5_L revealing six spatially distinct microenvironments (**Figure 8F-G**): with cluster-specific signatures (**Supplementary Figure 7A and Supplementary Table 7)** Finally, we examined the deconvolved topic composition for each of the six keratinocyte microenvironments (**Figure 8I and Supplementary Table 8**, Wilcoxon, padj<0.05) and confirmed by spatial mapping that each reflected distinct neighbourhood (**Supplementary Figure 7B**). Together, this analysis demonstrates that the P5_L, a 1-month-old *L. tropica* infection represents a complex inflammatory hyperplasia characterized by six spatially distinct epidermal microenvironments, each with unique molecular signatures and predicted cellular compositions.

## Discussion

In this study, we applied spatial transcriptomics to paired lesional and non-lesional skin biopsies from five Ethiopian patients with CL to define the cellular and molecular landscape of lesional tissue. While spatial transcriptomics has been applied to experimental human *L. major* infection ^33^ and *L. donovani*-associated CL in Sri Lanka ^34,35^, these studies have not investigated inter-patient heterogeneity. Leveraging the diverse Ethiopian CL clinical spectrum including infections with both *L. aethiopica* and *L. tropica* in our cohort, we discovered five distinct tissue-level pathological immune programs with differences in lesional cellular composition and spatial mapping. This also revealed patterns aligning with clinical presentations and disease stage: P1 and P5 showed neutrophil enrichment consistent with super-infection; mucosal lesions (P1, P3) exhibited sebaceous gland and IgA plasma cell enrichment; and late-stage lesions P2 (26 months) and P4 (11 months) displayed higher proportion of tissue repair macrophages and fibroblast topics. P2 (longest duration, hyperpigmented), showed the highest proportion of Fib3 (*SFRP2*, *PODN*) and endothelial topics, ECM remodelling genes, matrix-degrading enzymes, and myofibroblast markers paralleling healing phase findings in other chronic infectious diseases ^36^ including leprosy reversal reactions ^37^. Importantly, clinical classification (MCL/DCL) showed limited correlation with dominant tissue programs in this small cohort, suggesting that molecular pathotypes may transcend traditional clinical categories. These distinct pathotypes may represent conserved tissue programs with broader relevance to CL globally.

The presence of TLS in P1 represents, to our knowledge, the first report of lymphoid neogenesis in CL. Immunohistochemistry confirmed characteristic TLS architecture comprising B cells, T cells and a coordinated spatial expression of *CXCL13*, germinal centre and fibroblastic reticular cell markers. TLS formation in chronic infections often correlates with sustained antigen presence and attempts at local immune control ^38^. Paradoxically, P1 exhibited the highest parasite burden among all patients despite high cytotoxicity scores and elevated interferon-stimulated genes in TLS-adjacent regions (**Supplementary Figure 3E**). One explanation for this finding is the co-localisation of regulatory immune checkpoints (*IDO1*, *LAG3*) which may modulate cytotoxic effector responses. Our data suggest the need for further exploration of the role of TLS in CL pathogenesis, to determine whether they contribute to host protection or are associated with exaggerated immunopathology and chronicity.

Organized granulomas were prominent in P3 and P4, characterized by epithelioid macrophage cores enriched for lipid-associated macrophages and surrounded by lymphocytes. These macrophages expressed *CHIT1* and *CHI3L1*, consistent with lesional macrophages observed in *L. donovani* CL ^35^ and foamy macrophage phenotypes described in multiple sclerosis^39^ and coronary heart disease^40^. The accumulation of lipid-associated macrophages is a hallmark of chronic infectious granulomatous diseases, reflecting sustained phagocytic activity and lipid metabolism dysregulation ^41,42^.In tuberculosis, similar lipid-associated macrophage populations reside on granuloma boundaries ^43^ and may serve as niches for pathogen persistence ^44^.

Granuloma formation progresses through distinct phases:initiation, accumulation, effector, and resolution,each with characteristic cellular compositions and functions ^45^. Our spatial analysis revealed internal zonation in P4 granulomas with functional specialization including fibrotic zones, lipid-macrophage zones, T cell-DC interaction zones, and plasma-cytotoxic zones, consistent with spatial studies in microbial infection ^43^, Critically, the surrounding tissue microenvironments of P3 and P4 granulomas suggest they may represent different evolutionary phases: P3 granulomas were embedded in a proinflammatory milieu characterized by high MHC-II expression, class-switched immunoglobulins, and plasma cell infiltration, consistent with accumulation and effector phases. In contrast, P4 granulomas were embedded within an M2-polarized wound healing microenvironment, with elevated complement activation, distinct chemokine expression patterns and fibroblast markers. The organised zonation observed in P4, with *CXCL9* and *CCL19* upregulation, suggest a later stage of granuloma evolution as observed in cutaneous sarcoidosis ^46^ and healing phase granuloma in *L. braziliensis*-infected macaque models where fibroblasts proliferate at the granuloma periphery and invade granuloma structures^47^.

Our observation of granuloma heterogeneity parallels findings in infections by other intracellular pathogens such as *Mycobacteria tuberculosis*, where pulmonary granulomas enriched for B cells and fibroblasts differ substantially from visceral peritoneal omental granulomas dominated by myeloid cells ^48^, and in lung granulomas in macaques where different cellular compositions reflect high burden early granulomas (plasma cells, mast cells) versus low burden late (cytotoxic T cells) granulomas ^42^. The presence of organized and unorganised granulomas have been associated with in *L. aethiopica* lesions ^24,49^ but our analysis is the first to describe their molecular heterogeneity.

In P5, epithelial hyperplasia emerged as the most prominent and enriched pathological feature when compared to other patients. Inflamed keratinocyte signatures comprised 82% of total keratinocyte abundance across all epidermal layers, with widespread expression of interferon-stimulated genes, stress keratins, and antimicrobial alarmins. Six spatially distinct keratinocyte microenvironments were identified. Enrichment of inflamed keratinocytes with neutrophil recruitment is a well-characterized feature of leishmaniasis pathogenesis ^52^. Importantly, while epithelial inflammation was the dominant feature of P5, substantial dermal immune infiltration was also present, consistent with previous histologic report of *L. tropica* infection, illustrating that multiple active tissue programs likely co-exist within individual lesions. Whether the prominent epithelial response in this patient reflects *L. tropica*-specific biology^50^, early disease kinetics, or both, remains to be determined.

Recent histologic descriptions of *L.aethiopica* infection^24^ from 26 patients(18 LCL, 8 MCL) showed epidermal changes, organized granulomas in 3-12 month lesions and patchy inflammatory infiltrates in chronic lesions >12 months. We observed organized granulomas in P3 (MCL, 4 months) and P4 (LCL, 11 months), consistent with this temporal pattern. Notably, while the study associated organised granulomas with LCL and necrosis/ giant cells with MCL, our observation of organized granulomas in both clinical forms reinforce that molecular pathotypes transcend clinical classification. P2 (LCL, 26 months), with high stromal topic contribution, may represent the chronic healing phase while P5 (LCL) showed epidermal changes and inflammatory keratinocyte enrichment. Critically, our spatial approach dissects the molecular architecture underlying these histopathological patterns, revealing the distinct tissue programs and cellular associations that might define the tissue level heterogeneity in CL rather than the clinical form.

This exploratory study has the following limitations. The cross-sectional design cannot distinguish temporal progression from discrete pathotypes. The small sample size (n=5), single biopsies from index lesions that may not capture the entire lesional landscape and incomplete *Leishmania* species identification limits generalizability. Additionally, Visium spatial resolution does not achieve single-cell resolution, and reference-free deconvolution lacks single-cell validation. The absence of parasite spatial overlay precludes targeted host-parasite interaction analyses. Future studies with larger, longitudinal cohorts should integrate high resolution spatial parasite and host immune mapping to determine how parasite variation and immune programs drive disease progression, treatment failure, and clinical heterogeneity.

In conclusion, our study provides new insights into the complexity of the immunopathological response in patients with CL in Ethiopia. The identification of five distinct pathotypes, independent of clinical classification reveals that tissue-level immune programs define disease heterogeneity. By spatially dissecting granuloma evolution, cellular compositions, and stromal and epithelial remodelling programs, we establish a molecular framework for understanding CL pathogenesis that bridges descriptive histopathology with molecular immunology. These findings highlight the use of tissue biomarkers for guiding patient stratification and personalised immunomodulatory strategies tailored to distinct tissue programs in CL.

## Methods

### Ethics

The study protocol was approved by the Ethiopian National Research Ethics Review Committee (NRERC; 02/246/574/22), the University of Gondar Institutional Review Board (VP/RTT/05/156/2021), the Institute of Tropical Medicine Antwerp Institutional Review Board (1451/20), the Antwerp University Hospital Ethics Committee (UZA 624), and the Ethical Review Board of the Hull York Medical School (21-22.24). The study followed the Helsinki Declaration, Good Clinical Practices and local regulations. Written informed consent was obtained from all participants (or from the parent/guardian for adolescents). The study is registered at ClinicalTrials.gov (NCT05332093; registered 8 March 2022).

### Patient recruitment and sample collection

We enrolled six participants from the Spatial CL study investigating host-parasite interactions in Ethiopian CL at the Leishmaniasis Research and Treatment Centre (LRTC), University Hospital of Gondar (Gondar, Amhara Province, Ethiopia) between March-May 2022 with follow-up until November 2022. Participants were selected to represent diverse clinical presentations: localized cutaneous leishmaniasis (LCL, n=2), mucocutaneous leishmaniasis (MCL, n=2), and diffuse cutaneous leishmaniasis (DCL, n=2). Inclusion and exclusion criteria are available at Clinicaltrials.gov. Leishmania infection was verified by microscopy of Giemsa-stained skin slits and kDNA PCR of skin slit smears. Parasite genotyping by ITS-1 high-resolution melt PCR ^9^ and SureSelect ^51^ identified two *L. aethiopica* (P1, P2) and one *L. tropica* (P5) infection. Following clinical reassessment, one patient initially classified as DCL was diagnosed with leprosy and excluded; the remaining five patients (P1-P5) included two with mucosal involvement (P1, P3) and three with purely cutaneous lesions on the arm (P2), neck (P4), and leg (P5). Clinical data was collected and recorded in a REDCap database.

After enrolment, 4mm and 2mm punch biopsies (KAI Medical, Solingen, Germany) were collected from lesional (index lesion) and non-lesional skin (upper thigh or shoulder), respectively, snap-frozen in liquid nitrogen within 15 minutes, and stored frozen until Visium processing (10X Genomics, CA, USA).

### Tissue processing and assessment of RNA quality

Frozen tissue blocks were sectioned at 10 μm thickness and initial sections from each sample was assessed for RNA quality (RNA Integrity Number, RIN; 2.2-8.7). Due to low RIN values, we adapted the 10x Genomics Visium FFPE protocol for frozen sections^52^. Sections were fixed with 4% methanol-free formaldehyde (Thermofisher, Catalog number 28906) for 10 min at room temperature, washed, Hematoxylin and Eosin (H&E) stained and imaged as per Visium guidelines. Post imaging sections were de-crosslinked with a single wash of 100 μl 0.1 N HCl for 1 min. Subsequent steps followed standard Visium FFPE protocols to ensure probe-based transcript capture.

### Spatial transcriptomics

Following deparaffinization, hybridization, extension, and ligation steps, cDNA libraries were constructed using the Visium Spatial Gene Expression reagent kit (10x Genomics) adapted for partially fixed frozen sections. Sequencing was performed on an Illumina NovaSeq 6000 instrument. Raw FASTQ files were processed using SpaceRanger (10x Genomics) to align reads to GRCh38 and generate feature-barcode matrices. Quality control metrics (total UMI counts, gene counts per spot) were assessed to filter out low-quality spots.

### Data integration, normalization, and clustering

Filtered gene-barcode matrices were loaded into Seurat (Seurat_4.3.0, R 4.2.3) and normalized using SCTransform (default parameters). Data were integrated across samples using integration anchors. Spots with <100 total and genes with <10 reads across all spots were removed. Principal component analysis (PCA) was performed on the integrated data, and clusters were identified using the Louvain algorithm at a suitable resolution. UMAP was used for two-dimensional visualization of spatial domains. Differential gene expression (DGE) between clusters and conditions (lesional vs non-lesional, and patient specific) was performed using a Wilcoxon rank sum test with (minimum detection fraction >0.1, fold change >0.5) with Bonferroni correction.

### Deconvolution of Visium spots and visualization

Cell type abundance in each Visium spot was estimated using STdeconvolve (v1.3.2 ^29^) a reference-free deconvolution tool using latent Dirichlet allocation to identify recurrent gene expression patterns (“topics”) representing putative cell types or states. The output provides gene expression profiles per topic and topic proportions per spot. Some topics captured mixed cellular signatures reflecting spatial co-localization or coordinated functional programs. Following quality control filtering, keratin genes (KRT*) and immunoglobulin genes (IGH*, IGL*, IGK*) were excluded as their high expression levels obscured other immune cell populations during deconvolution. However, *IGHM* and *IGHA1* were reintroduced to discriminate non-class-switched (IgM+) and class-switched (IgA+) plasma cells/B cells. Overdispersed genes were identified using the restrictCorpus() function (removeAbove = 0.95, removeBelow = 0.01, alpha = 0.05) to capture genes with high biological variability. To ensure adequate T cell subset representation, we supplemented the over dispersed (OD) gene set with curated T cell markers. The final gene combined all OD genes with immune markers present in the dataset. Deconvolution was performed with k=26 topics for the combined non-lesion and lesion dataset and k=50 topics for the lesion-only dataset. Topics with similar expression profiles were subsequently merged for final annotations.

### Paired analysis of lesional vs. non-lesional tissue

Cell type composition differences between paired lesional and non-lesional samples were assessed using linear mixed-effects models (lme4 v1.1-35, lmerTest v3.1-3) on spot-level deconvolution data (n=5,562 spots, 5 patients). For each of 23 topics, we fitted: Proportion ∼ Condition + (1|Patient_ID), where Condition (lesional vs non-lesional) was a fixed effect and Patient_ID a random intercept to account for paired samples and patient-specific baselines. Models were fitted using restricted maximum likelihood (REML). From each model, we extracted: (1) fixed effect coefficient (β) representing mean difference between conditions, (2) standard error, t-statistic, degrees of freedom, and p-value, (3) Cohen’s d as standardized effect size (β / √[σ²_patient + σ²_residual]), (4) fold change (lesional/non-lesional mean proportions), and (5) intraclass correlation coefficient (ICC = σ²_patient / [σ²_patient + σ²_residual]) quantifying variance attributable to between-patient differences. P-values were adjusted using Benjamini-Hochberg FDR correction. Cell types with padj < 0.05 and fold change >1.5 or <0.67 were classified as significantly enriched in lesional or non-lesional tissue, respectively. Effect sizes were interpreted per Cohen’s conventions: |d| < 0.2 (negligible), 0.2-0.5 (small), 0.5-0.8 (medium), >0.8 (large).

### Module score calculation

Gene signature module scores for cytotoxicity (*GZMB, PRF1, GNLY, NKG7*) and fibrosis (*COL1A1, COL1A2, COL3A1, COL6A1, COL6A2, MMP2, TIMP2, TIMP3, ACTA2, PDGFRB, SPARC, FBLN1, FBLN2, DCN*) were calculated using Seurat’s AddModuleScore() with default parameters. To identify regions with the highest cytotoxic activity, spatial spots were stratified based on the cytotoxicity module score distribution. High cytotoxic spots were defined as scores ≥Q3 (top 25%); low cytotoxic spots comprised the remaining 75% (score <Q3).

### Spatial Annotation and Region Classification

Spatial regions were manually annotated using 10x Genomics Loupe Browser based on underlying H&E histology and immune infiltration patterns. For both P3 and P4 lesions, granulomatous regions were delineated by characteristic histological features (epithelioid histiocyte aggregates) and CD68/ CD8A immunofluorescence staining. For P4, regions were further classified into four categories: organized immune structures (characterized by dense, structured lymphoid aggregates), diffuse immune infiltrates (scattered immune cells near granulomas), uninvolved tissue (minimal immune presence), and remaining tissue.

### Cell Type Enrichment Analysis

Cell type enrichment was assessed by comparing deconvolved topic proportions between regions or conditions. For pairwise comparisons, two-sided Wilcoxon rank-sum tests were performed (minimum 5 spots per group). For ≥three categories, Kruskal-Wallis tests were followed by pairwise Wilcoxon tests. Fold enrichment was calculated as the ratio of mean proportions between groups. P-values were adjusted using Benjamini-Hochberg correction (padj < 0.05 considered significant).

### Quantification of keratinocyte inflammation across patient samples

The inflamed keratinocyte ratio was calculated per spot as the sum of inflamed keratinocyte topic proportions (inf_BasalKer + inf_SupraBasal + inf_TopKer) divided by the sum of all keratinocyte proportions (inflamed + non-inflamed types). Non-inflamed keratinocyte topics included BasalKer, SuprabasalKer, LC_basal and TopKer.

### Functional annotation and pathway enrichment analysis

DEG lists from each comparison were subjected to functional annotation using Reactome databases using StringDB (STRING 12.5 ^53^). Significantly enriched pathways were identified (adjusted p-value < 0.05) and visualized using dot plots as exported from StringDB with increasing signal metric (weighted harmonic mean of observed/expected ratio and –log (FDR)). All terms were grouped together with similarity score of 0.8 (based on Jaccard index).

*Subclustering of organized immune regions in P4 and lesional keratinocytes in P5* Dimensionality reduction was performed using UMAP, and clusters were identified using Louvain clustering (10 PCs and resolution 1-1.2). Marker genes per subcluster were identified using Wilcoxon rank-sum tests comparing each cluster against all others.

### Immunofluoresence (IF)

For IF, 10 μm sections were cut from frozen blocks and mounted on SuperFrost Gold slides (11976299). Sections were fixed in 4% paraformaldehyde for 30 minutes and followed by 0.3% Triton X 100 mediated permeabilization for 15 minutes. Slides were blocked with normal serum for 1 hour at room temperature (RT). For multiplex IF staining, sections were incubated with unconjugated primary antibodies: CD68 (KP1, ab955, abcam, 1:200) for myeloid cells, and CD8a (C8/144B, 372904, Biolegend, 1:100, AF594) for T cells sequentially for 1hr at RT. For separate experiment, sections were stained with CD20 (IGEL/773, NBP2-47840C, Novus Biologicals, 1:100, Dylight-650) for B cells and CD3E (CD3-12, ab11089, Abcam, 1:300]) for T cells.

For parasite detection, sections were incubated with unconjugated anti-OpB antibody (*Leishmania* OPB (10 μg/ ml, provided by Jeremy Mottram, University of York) overnight at 4^0^C. Following primary antibody incubations, sections were washed and incubated with appropriate fluorophore-conjugated secondary antibodies (Donkey anti-sheep AF647 catalogue# A21448, Invitrogen for OPB, IgG Donkey anti-rat AF555, ab150154, Abcam for CD3E and CF750 Goat Anti-Mouse IgG, Generon, 20463-250uL for CD68 at 1:500 for 30 minutes at RT. Nuclei were counterstained with YOYO1 (0.2µM) for 1hr. Slides were mounted with Prolong Gold. IF images were acquired using a Zeiss, Axio Scan.Z1 slide scanner at 10x. Images were visualized and analyzed using ZEN Microscopy Software.

### Statistical Testing and Visualization

Unless otherwise specified, all tests were two-sided. For multiple pairwise comparisons, Benjamini-Hochberg correction was applied to control FDR. Box plots display median (center horizontal line), interquartile range (box), and 1.5× IQR whiskers. Violin plots show kernel density estimates of the full distribution. In dot plots, size of the dot shows percent expressed in the region/ cluster and color of the dot indicates scaled expression. Statistical significance is indicated as: *p < 0.05, **p < 0.01, ***p < 0.001, ****p < 0.0001. All analyses were performed in R (version 4.5.0) using Seurat (version Seurat_5.3.0), STdeconvolve (version 1.3.2), SCpubr (version 3.0.0), ComplexHeatmap (version 2.24.1), Scatterpie (v0.2.6), ggpubr (v0.6.0) and ggplot2 (version 3.5.2) packages.

## Data and materials availability

The sequencing data have been deposited in GEO (GSE317987). The processed data are available as Rds files on Zenodo: https://doi.org/10.5281/zenodo.18471139. Analysis code is available at https://github.com/NidhiSDey/SpatialCL.git.

## Author contributions

NSD, TTP, JVG, WA and PMK: conceptualization.

SPATIAL CL CONSORTIUM, NSD, TTP, SD, MK, TM, HF, PM, MAD, JCD, WA, PMK: Investigation, methodology

NSD, TTP, SD, WA, JCD, JVG, PMK: writing, review and editing,

NSD: Data curation, Validation, Formal analysis, Visualization, Writing-original draft SPATIAL CL CONSORTIUM, TTP, JVG, PM, MAD, JCD, MSA, MW, PMK and WA: Project administration, resources

WA, PMK, MW: supervision

WA, JVG, PMK: funding acquisition

## Acknowledgments

This work was supported by funding from the Dioraphte Foundation (200200401 to WA, JVG, PMK), and Wellcome Trust Investigator Award (WT224290 to PMK). TP received a post-doctoral Fellowship from the VALIDATE Network and a mobility grant from the Research Foundation Flanders (FWO – K224622N). The authors thank all patients and their families who took part in this study and all members of the Kaye laboratory at Hull York Medical School, University of York for their useful comments and suggestions on this project. Biorender.com was used to generate the graphical abstract.

---

**Supplementary Figure 1.**
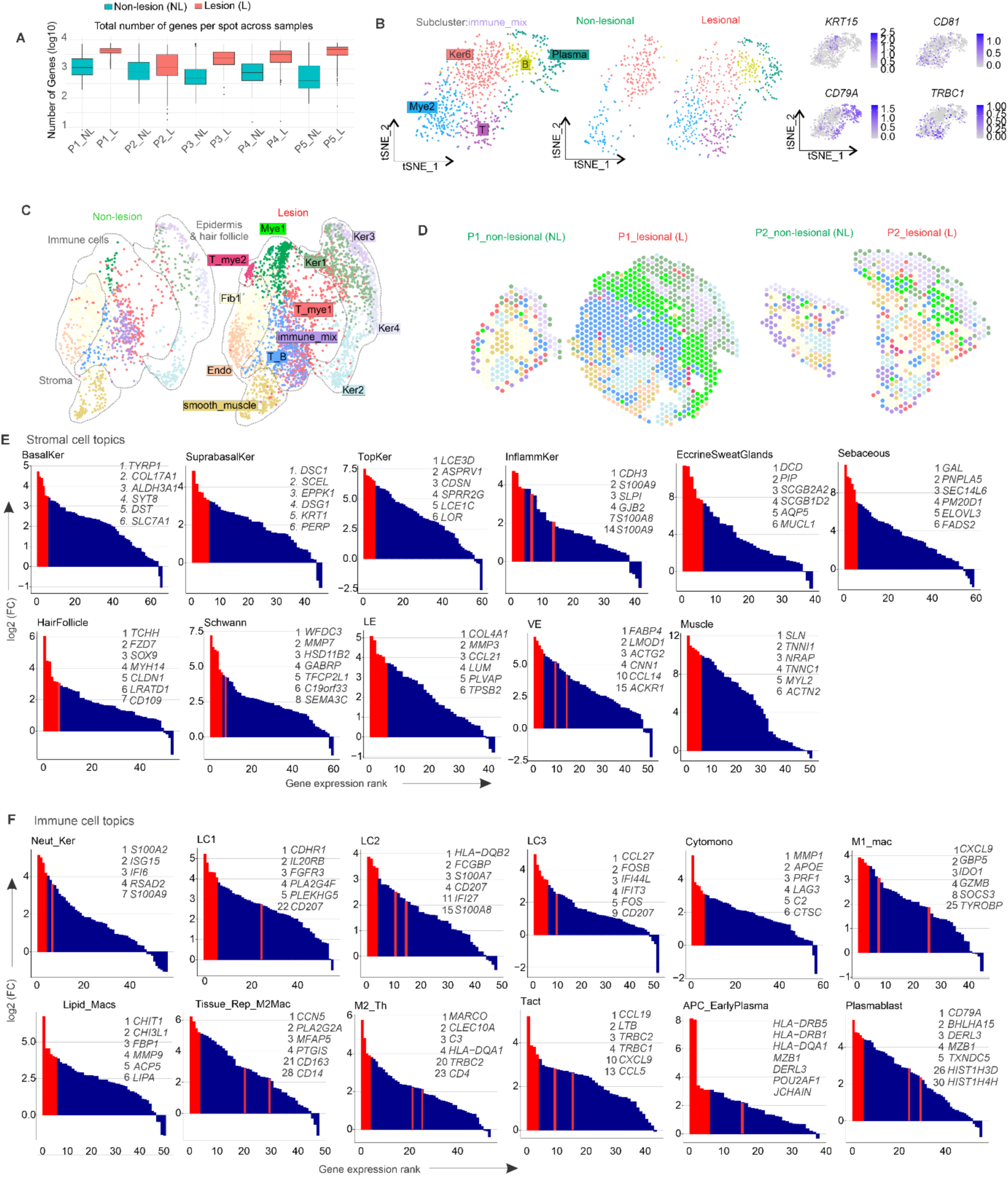
Quality control metrics and cell type analysis in paired non-lesional and lesional skin of CL patients. **A**, Number of detected genes (nFeature metric; log10) per spot across non-lesional (NL) and lesional (L) samples (P1-P5). **B**, UMAP showing subclusters of cluster immune_mix from **Figure 2A**: immune rich: (Mye2, T, B, Plasma) and keratinocyte rich (Ker6). Right panel UMAP is split by L and NL tissue. Left panel is immune_mix subcluster feature plot showing enrichment of *KRT15*, *CD81*, *CD79A*, *TRBC1* in Ker6, Mye2, B, Plasma subdomains. **C**, UMAP from Figure1A split by L and NL skin. **D**, Spatial plots from two representative patients P1 and P2: L and NL showing skin domains from **Figure1A**. **E-F**, Genes expression in each STdeconvolve computed stromal (**E**) and immune (**F**) deconvoluted topic in NL and L combined dataset.

**Supplementary Figure 2:**
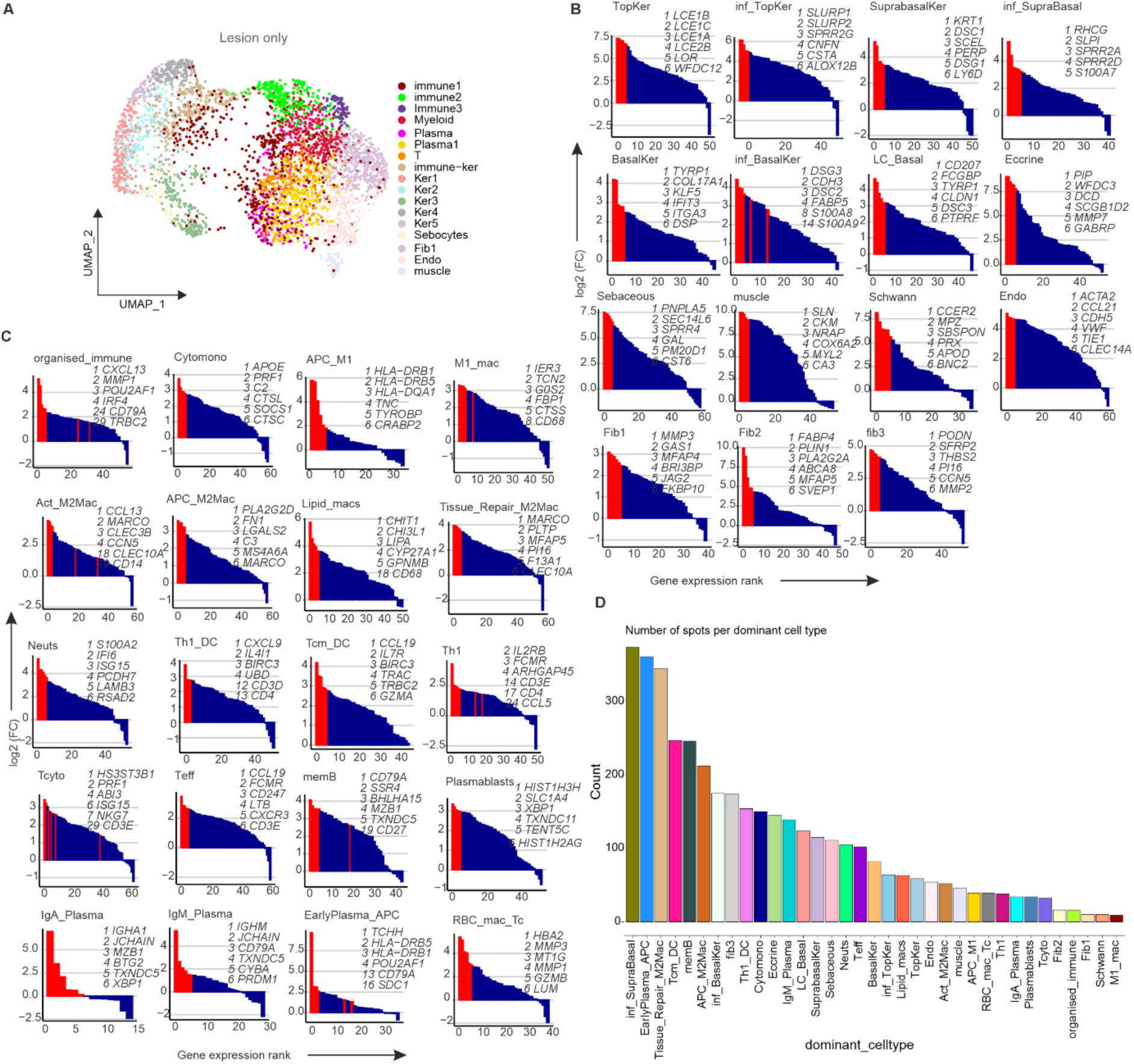
Gene expression and spot counts of deconvoluted cell types in all patients’ L skin samples. **A**, UMAP of all lesional skin spots colored by cluster identity, imputed on top marker genes. **B-C**, Top 6 genes expressed in each cell type from subsetted L skin (n=5) from stromal (**B**) and immune (**C**) deconvoluted topics. **D**, Bar chart showing spot count per dominant topic.

**Supplementary Figure 3:**
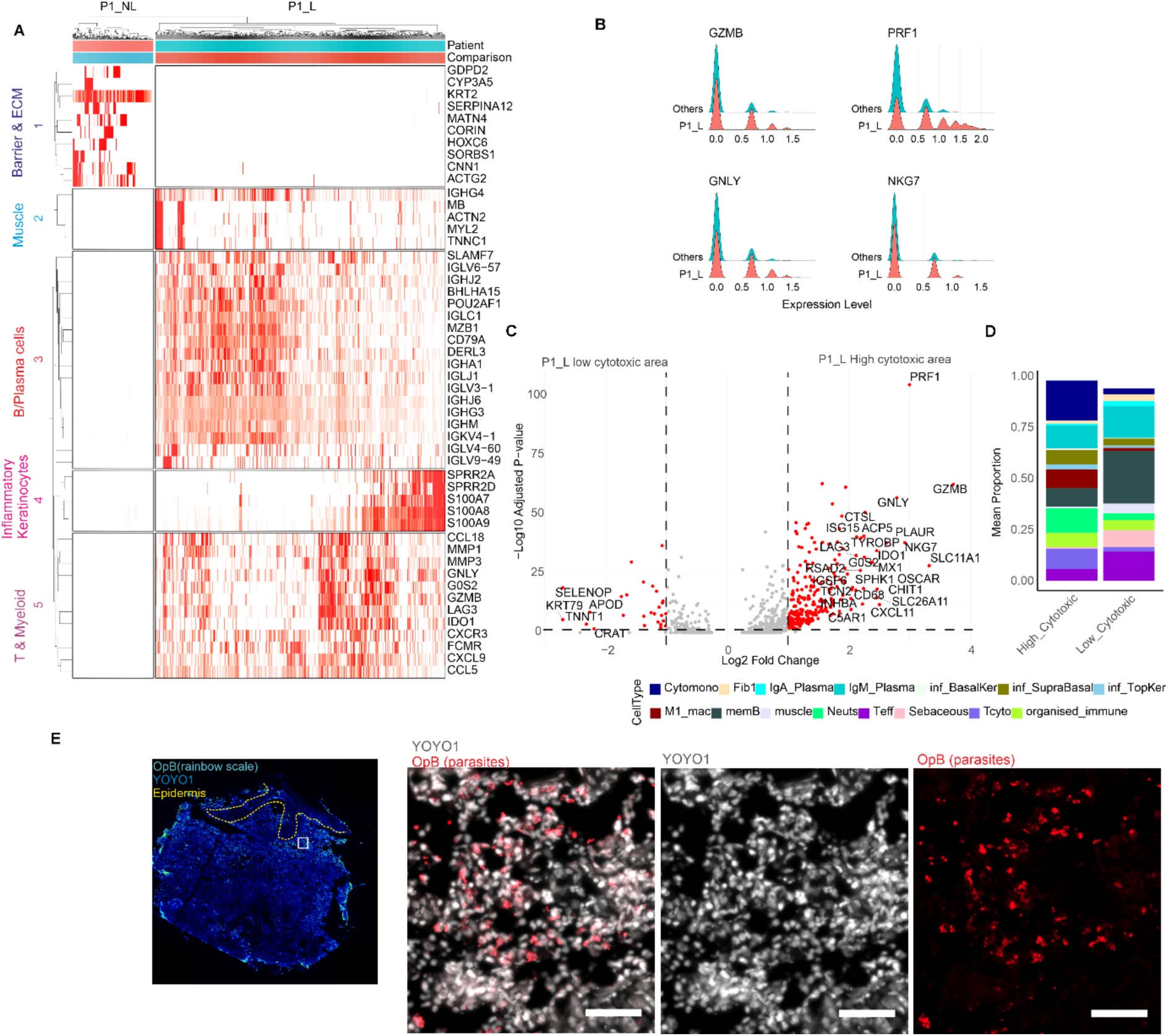
Transcriptional and cellular characterization of cytotoxic regions in P1 lesional skin. **A**. Scaled expression heatmap of DEGs between P1 non-lesional (NL) and lesional (L) skin with functional annotation. **B**, Ridge plot comparing cytotoxicity marker expression between P1_L and other lesional samples (P2-P5_L). **C**, Volcano plot showing differentially expressed genes between high and low cytotoxicity spots in P1_L; red: significant genes (adjusted p < 0.05); top 30 labeled. **D**, Relative proportions of deconvolved cell types in high versus low cytotoxic regions of P1_L (top 15 cell types enriched in high cytotoxic areas). **E,** IHC staining for leishmania parasites (OPB: Oligopeptidase B) in P1_L lesion, showing parasites in rainbow scale on whole image (left most panel; nuclei: YOYO1: blue; scale bar=100µm) and in magnified regions from it (nuclei: YOYO1: grey; scale bar: 20µm).

**Supplementary Figure 4.**
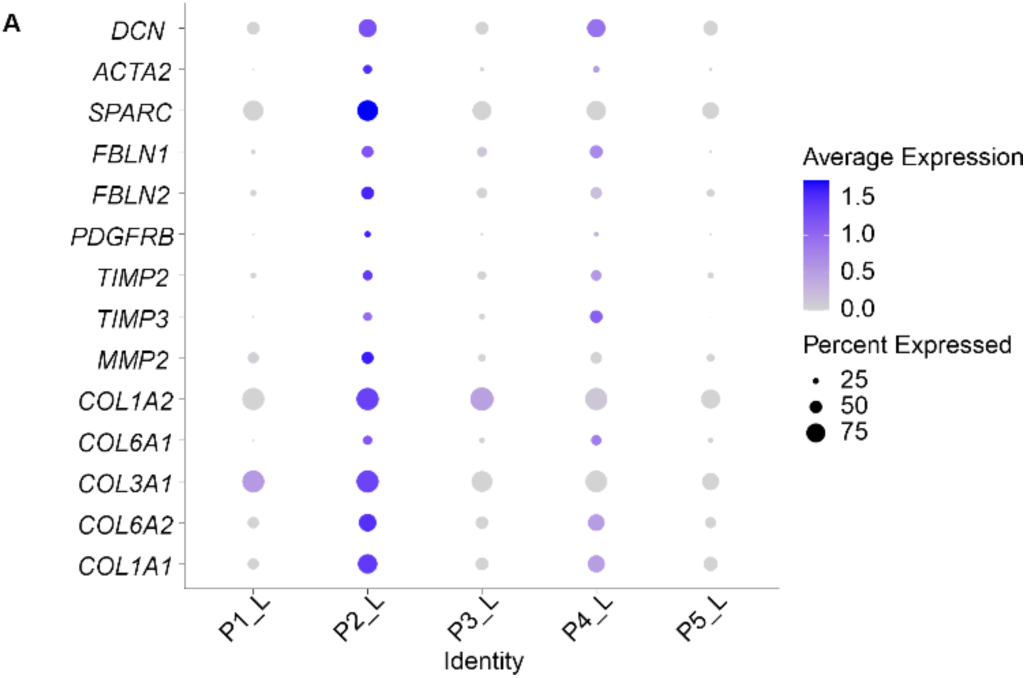
Expression of lesional fibrotic signature genes across patients. **A**, Dot plot showing expression of genes comprising the fibrotic score module across all lesional samples (P1-P5 L).

**Supplementary Figure 5:**
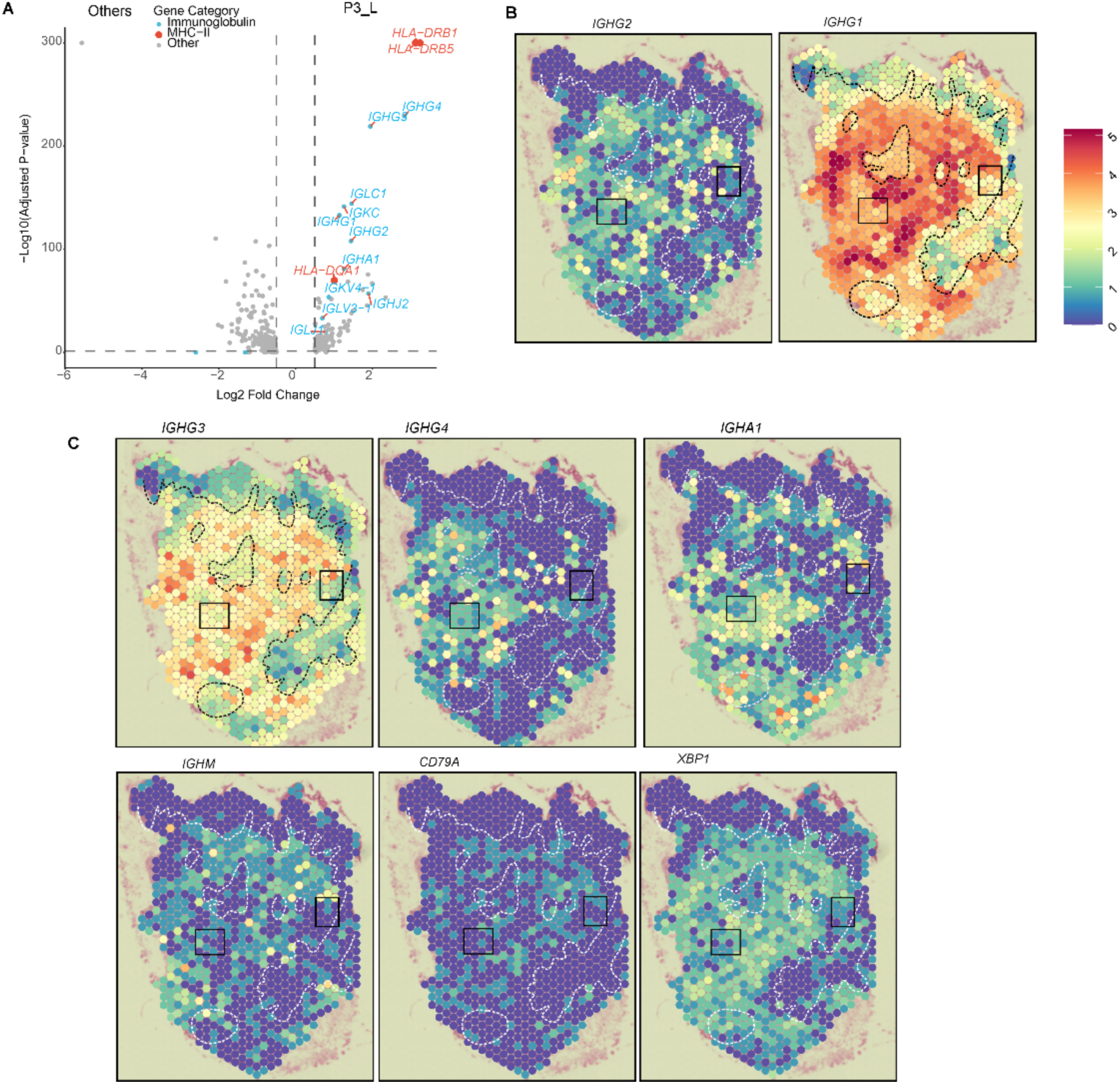
Immunoglobulin and MHC class-II gene expression in lesional skin from P5. **A**, **A**, Volcano plot showing DEGs between P3_L and other lesional samples (P1-2 L, & P4-5_L). Antigen presenting markers in red (log2FC > 0.5, padj < 0.05, Wilcoxon); immunoglobulin genes in blue. **B-C**, Spatial expression heatmaps showing distribution of B/ plasma cell associated genes in P3_L skin (*IGHG1-4*, *IGHA1*, *IGHM*, *CD79A*, *XBP1*). Regions 1 and 2 from **Figure 6C** are indicated by black rectangles.

**Supplementary Figure 6.**
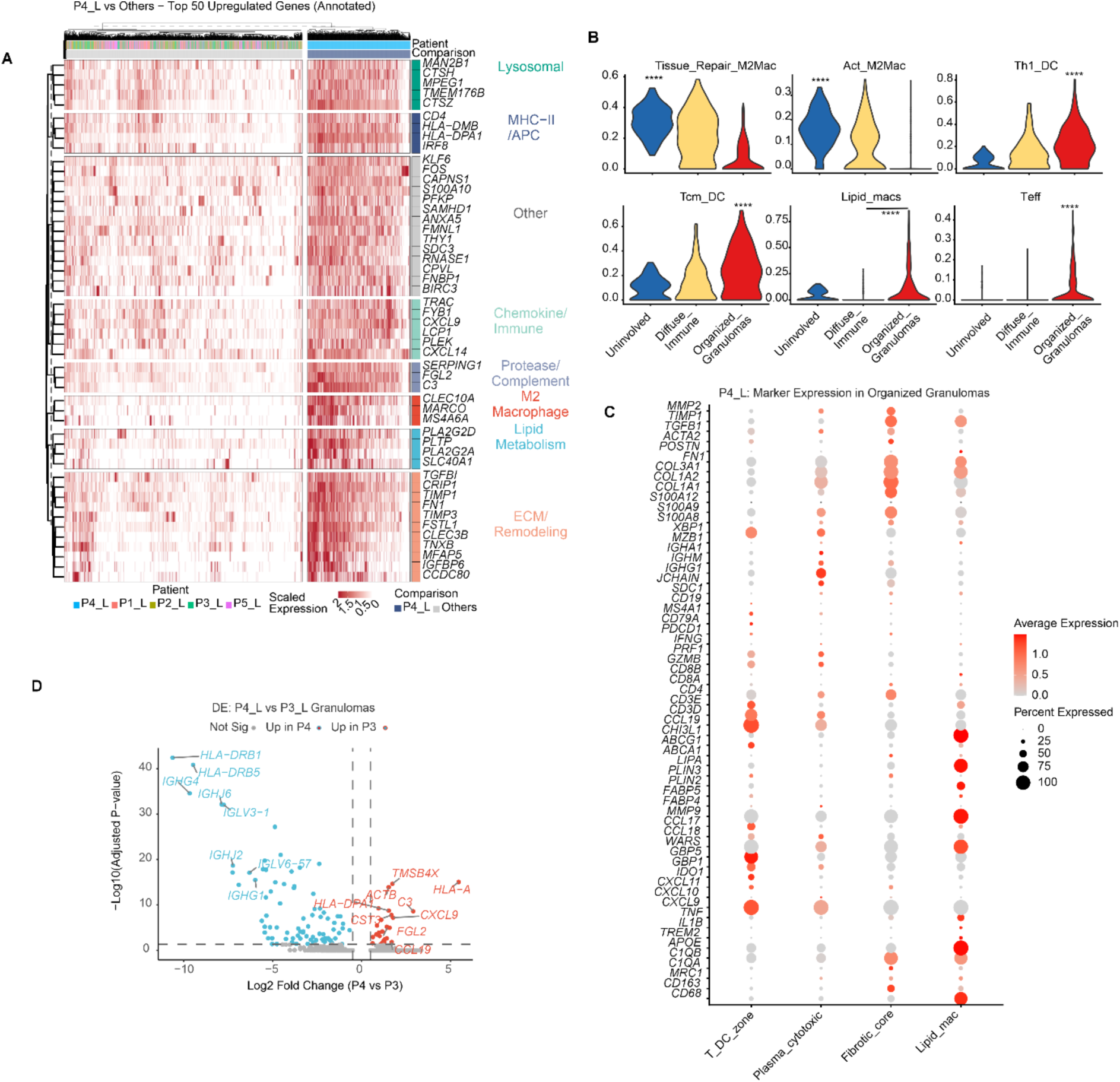
Differential expression of markers in P4 L microenvironment. **A**, Heatmap showing scaled expression of the top 50 upregulated genes (padj < 0.05, log2FC > ±0.25; Wilcoxon) between patient P4 L and all other patient samples (P1-P3 and P5 L) with functional annotation. **B**, Violin plots of top 6 deconvoluted topics in categories from Figure 6E. **C**, Dot plot of selected markers in subclusters of organised granuloma spots. **D**, DEGs between manually annotated P4_L and P3_L granulomas samples (volcano plot, padj<0.05, log2FC>0.5, top genes labelled).

**Supplementary Figure 7.**
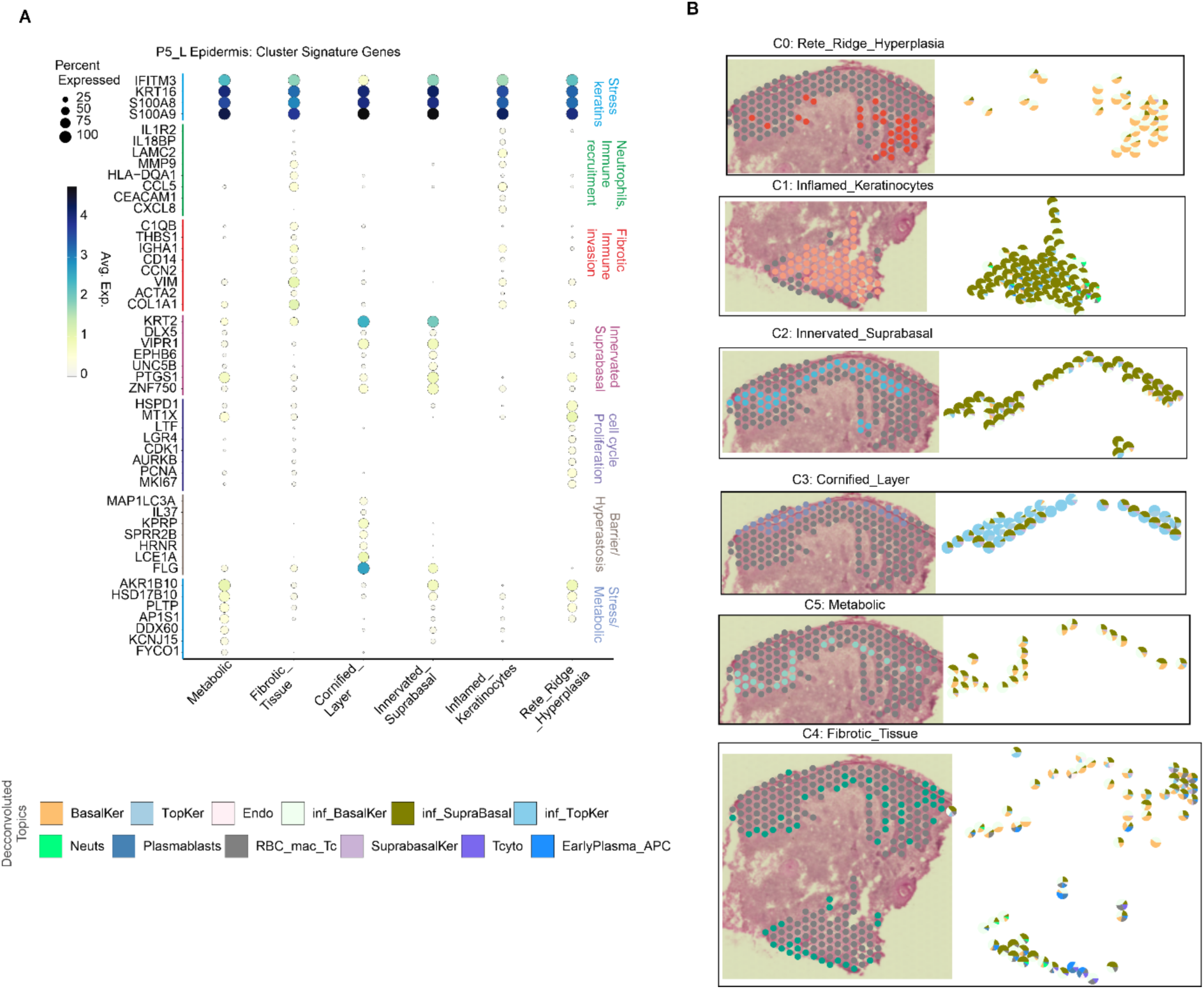
Gene expression and spatial distribution of epidermal clusters in P5_L. **A**, Dotplot of cluster-specific signature genes from **Figure 8F**. Genes categorised by biological function. **B,** Spatial scatterpie charts showing topic composition for individual spots within each epidermal subcluster.

