## Supplemental Table 1 for "Spatial mapping of Ethiopian cutaneous leishmaniasis lesions reveals distinct tissue level immune programs"

**Table 1. Sociodemographic data of patients**

| | |
|---|---|
| Age (years)* | 21.5 (20.3-25) |
| Sex (Male:Female) | 6:00 |
| Occupation | |
| *Student* | 2/6 (33.3%) |
| *Farmer* | 2/6 (33.3%) |
| *Clerk* | 1/6 (16.7%) |
| *Unemployed* | 1/6 (16.7%) |
| Level of education | |
| *Primary school* | 1/6 (16.7%) |
| *High school* | 4/6 (66.7%) |
| *None* | 1/6 (16.7%) |

*values given in Median (Interquartile Range)
