## Supplemental Text 1 for "Spatial mapping of Ethiopian cutaneous leishmaniasis lesions reveals distinct tissue level immune programs"

**Detailed description of the Spatial CL Consortium**

This African-European consortium was created to investigate the underlying host-parasite interactions in Ethiopian patients with different clinical presentations of cutaneous leishmaniasis. The study includes an in-depth characterization of the infecting parasite, host metabolic status and associated host immune response in a spatial and single-cell resolved manner. The clinical study has been registered on Clinicaltrials.gov (NCT05332093) and consortium members are listed below:

- Leishmaniasis Research and Treatment Centre, University of Gondar, Ethiopia:
    - Dr. Mezgebu Silamsaw Asrem
    - Dr. Eleni Ayele
    - Dr. Helina Fikre
    - Dr. Yonael Mulat
    - Tigist Mekonnen
    - Mekibib Kassa
    - Tadfe Bogale
    - Asnakew Engidaw Mereed
    - Tadele Mulaw
    - Roma Melkamu
    - Arega Yeshanew
    - Aman Mossa
    - Zemeney Mulugeta
    - Dilargachew Dessie Dawit
    - Aschalew Tamiru
    - Desalegn Adane
    - Saba Atnafu
    - Jemal Yasin

- Department of Immunology and Molecular Biology, University of Gondar, Ethiopia:
    - Abiy Ayele Angelo
    - Yetemwork Aleka
    - Hana Yohannes
    - Helen Terefe

- Department of Dermatology and Venereology, University of Gondar, Ethiopia:
    - Dr. Mikias Woldetensay
    - Dr. Abebe Sinknew Seid
    - Dr. Mesfin Malede Nigussie
    - Dr. Anemut Leykun Ayele
    - Dr. Beza Tesfalem
    - Dr. Tesfaye Ketema
    - Dr. Mengistu Tilaye Belay
    - Dr. Seidu Biresaw

- Department of Surgery, University of Gondar, Ethiopia
    - Dr. Getachew Yenus Aman

- York Biomedical Research Institute, Hull York Medical School, United Kingdom:
    - Prof. Paul Kaye

- Dr. Nidhi Sharma Dey
- Dr. Shoumit Dey

- Multimodal Imaging Institute, University of Maastricht, The Netherlands:
    - Prof. Ron Heeren
    - Prof. Benjamin Baluff
    - Dr. Isabeau Vermeulen
    - Dr. Jianhua Cao

- Department of Plastic, Reconstructive and Aesthetic Surgery & Departement Antwerp Surgical Training, Anatomy and Research Centre, University Hospital of Antwerp, Belgium:
    - Prof. Filip Thiessen

- Molecular Parasitology unit, Institute of Tropical Medicine, Belgium:
    - Prof. Jean-Claude Dujardin

- Experimental Parasitology unit, Institute of Tropical Medicine, Belgium:
    - Prof. Malgorzata Domagalska
    - Kaoutar Choukri

- Trypanosoma unit, Institute of Tropical Medicine, Belgium:
    - Dr. Pieter Monsieurs

- Unit of Neglected Tropical Diseases, Institute of Tropical Medicine, Belgium:
    - Prof. Johan van Griensven
    - Dr. Saskia Van Henten
    - Dr. Myrthe Pareyn

- Clinical Immunology unit, Institute of Tropical Medicine, Belgium
    - Prof. Wim Adriaensen
    - Dr. Thao-Thy Pham
    - Nicky de Vrij
    - Anke Van Hul
